# Sociodemographic Disparities in Tafamidis Initiation and Clinical Outcomes in ATTR-CM Across the United States

**DOI:** 10.64898/2026.06.12.26355533

**Authors:** Nicole Cyrille-Superville, Hanna K. Gaggin, Andrew M. Rosen, Margarita Udall, Liana Hennum, Bret Zeldow, Xingyu Gao, Elizabeth Nagelhout, Allison Keshishian, Margot K. Davis

## Abstract

**BACKGROUND:** Transthyretin amyloid cardiomyopathy (ATTR-CM) is a progressive, life-threatening disease. Sociodemographic factors may influence time to treatment initiation and resulting clinical outcomes, yet these relationships are poorly characterized.

**OBJECTIVE:** Assess the effects of sex and race on tafamidis initiation and subsequent outcomes and their interaction with factors such as ATTR-CM type and social deprivation measures.

**METHODS:** A retrospective cohort analysis was conducted using the US Komodo Healthcare Map^®^ (01/2016-06/2024) among patients with amyloidosis, identified by ICD-10-CM diagnosis codes. Cumulative incidence of treatment initiation and survival probabilities for cardiovascular-related hospitalization (CVH) or death were estimated by Kaplan-Meier, stratified by sex and race. Cox proportional hazards models were fitted for both endpoints to estimate hazard ratios, adjusting for demographics and clinical characteristics.

**RESULTS:** Of 11,311 patients identified, White and Black patients (n=9,223) were included in subsequent analyses. Within 12 months of diagnosis, White women had the lowest cumulative incidence of tafamidis initiation (11.4%), followed by Black women (22.0%), Black men (26.7%), and White men (31.0%). Event-free survival at 12 months was lowest in Black women (42.9%), followed by Black men (46.8%), White women (48.6%), and White men (54.4%). Median (95% CI) time to CVH or death was shortest for Black women (8.0 months [6.8-10.0]) followed by Black men (9.9 months [8.8-12.0]), White women (11.0 months [9.6-13.0]), and White men (15.0 months [14.0-16.0]).

**CONCLUSIONS:** In this large, real-world cohort of US patients with ATTR-CM, sex and race contributed to disparities in tafamidis initiation and survival, underscoring compounded disparities in both access and outcomes.

## INTRODUCTION

Transthyretin amyloid cardiomyopathy (ATTR-CM) is a progressive, life-threatening condition caused by cardiac deposition of pathogenic amyloid fibrils from misfolded transthyretin (TTR) protein.^1^ There are 2 recognized clinical forms of ATTR-CM: wild-type ATTR-CM (ATTRwt-CM) and variant ATTR-CM (ATTRv-CM). ATTRv-CM is caused by pathological mutations in the *TTR* gene. ATTR-CM predominantly affects older adults; however, patients with ATTRv-CM typically present at a younger age than patients with ATTRwt-CM.^1^ Despite advances in noninvasive imaging, growing disease awareness among clinicians, and the availability of disease-modifying treatments, ATTR-CM remains underdiagnosed, with diagnostic delays of up to 6 years after symptom onset.^1,2^ There are suggestions that disparities exist in diagnosis and time to treatment initiation, which may result in inequities in patient outcomes.^1,3^ It has been reported that Black patients present with more advanced disease at diagnosis compared with White patients, and women with ATTRv-CM are generally diagnosed approximately 7 years later than men with the same variants.^4,5^

Tafamidis was the first and only disease-modifying treatment approved for ATTR-CM at the time of the study period. Tafamidis is most effective when initiated early.^6^ Real-world analyses and long-term extension studies confirm that earlier treatment initiation results in more sustained clinical benefits, yet treatment access and delays remain inequitable.^7-10^ In particular, the high annual cost of tafamidis (listed at $277,709) poses a prohibitive burden for patients who are uninsured or underinsured, though patient assistance programs offer relief for those who can navigate this support.^11-14^ Overall, the benefit of ATTR-CM treatment observed in clinical trials does not always translate into real-world settings, with persistently high rates of morbidity and mortality in populations with compounded barriers in access to care.^9,10^

In addition to diagnostic delays, evidence suggests that sociodemographic factors, particularly sex and race, influence both time to treatment initiation and key clinical outcomes in ATTR-CM, including cardiovascular-related hospitalization (CVH) and all-cause mortality, yet these relationships are poorly characterized.^4,15^ The underrepresentation of these populations in pivotal trials limits our understanding; Black patients comprised only 14.3% of participants in the tafamidis phase 3 ATTR-ACT trial,^6,7^ yet they accounted for 2.4% of those with ATTRwt-CM and 51.9% of those with ATTRv-CM.^6,7,16^ Women have been similarly underrepresented in ATTR-CM trials, comprising only 8.7% of patients within the tafamidis arm.^6^ This is highly relevant in real-world clinical practice, given evidence that the true prevalence of disease in women has been underestimated. Some of the reasons for underdiagnosis in women may be due to the use of diagnostic criteria that incorporate a single threshold of left ventricular wall thickness for all individuals (ie, ≥12 mm), which may systematically miss female patients who generally have smaller hearts and therefore thinner ventricular walls.^17,18^ Differences between demographic groups in ATTRv-CM prevalence, age, disease stage at presentation, and disease phenotype suggest that universal treatment approaches may not address the specific needs of all patient populations.^17,18^ Using a large, real-world US cohort, we aimed to quantify sociodemographic disparities in time to tafamidis initiation and key clinical outcomes of CVH and all-cause mortality, hypothesizing that sex and race independently and synergistically determine treatment access and outcomes.

## METHODS

### STUDY DESIGN AND DATA SOURCE

A retrospective cohort study was conducted using the Komodo Healthcare Map^®^, a US longitudinal claims database covering medical and pharmacy claims across commercial and public payers. The observation window was January 1, 2016, through June 30, 2024, representing the period over which Komodo claims data were available. The first portion (2016-2019) served as a baseline look-back window requiring the absence of qualifying ATTR-CM diagnosis codes or tafamidis claims, thereby excluding prevalent ATTR-CM cases. The patient identification period was from May 3, 2019 (date of United States [US] Food and Drug Administration [FDA] approval of tafamidis for ATTR-CM), through June 30, 2024, representing the window for qualifying new ATTR-CM diagnoses had to occur for a patient with newly diagnosed ATTR-CM to enter the cohort. Thus, all included patients were diagnosed on or after the May 3, 2019, FDA approval date of tafamidis for ATTR-CM. The diagnosis index date was defined as the earliest occurrence of a qualifying ATTR-CM diagnosis code. The treatment index date was defined as the date of the initial tafamidis claim. Both the diagnosis and treatment index dates were required to occur during the patient identification period. Patients were followed until death, disenrollment, or end of the study period, whichever occurred first.

The Komodo Healthcare Map comprises medical and prescription claims data from several sources. It is characterized by proprietary partnership with >150 key national payers and consortiums, representing over 150 million payer lives. The database is representative of the US commercial-, Medicare-, and Medicaid-insured populations, and provides insights into the complete patient journey over time. The Komodo database complies with the Health Insurance Portability and Accountability Act. Given the anonymized, de-identified nature of the data, patient consent was not possible or required, and institutional review board approval to conduct this study was not necessary. Only aggregate cohort-level results were reported, without identification of any healthcare providers or healthcare facilities.

### STUDY POPULATION

Patients were eligible if they were ≥18 years of age and had ≥2 claims for amyloidosis (International Classification of Diseases, Tenth Revision, Clinical Modification [ICD-10-CM] codes E85.0, E85.1, E85.2, E85.4, E85.82) occurring on separate days, along with ≥2 claims for a cardiac-related ICD-10-CM code during the identification period. Patients were required to have ≥6 months of continuous enrollment before the diagnosis index date and ≥1 day of enrollment after the diagnosis index date. Patients were excluded if they had >1 claim (on separate days) for multiple myeloma (MM), light chain (AL) amyloidosis (E85.81), or hematopoietic stem cell transplantation any time during the study period, tafamidis use or ATTR-CM diagnosis any time before the diagnosis index date (to ensure a newly diagnosed population), or unknown sex.

### VARIABLES

The primary outcome was time from ATTR-CM diagnosis to tafamidis initiation. The secondary outcome was time from diagnosis to CVH or all-cause death. Diagnosis was defined as the earliest of the 2 amyloidosis ICD-10-CM diagnosis codes, and the treatment index date was defined as the first tafamidis claim. CVH was defined as inpatient encounters with a CV-related ICD-10-CM code in any position (**Supplemental Table 1**); all-cause death was identified using claims-based mortality flags.

Patient characteristics evaluated included age, sex, race (self-identified), year of diagnosis, US region, and insurance type. Socioeconomic status was measured by Social Deprivation Index (SDI), a composite measure of area-level deprivation as it relates to social determinants of health, including poverty, education, employment, housing, transportation, and other household characteristics.^19^ This metric is used to quantify levels of disadvantage and inequities in small areas of the US. SDI scores range from 1 to 100, with a higher score indicating higher social vulnerability. SDI was dichotomized as ≤80 (nondisadvantaged) vs >80 (disadvantaged) in line with previous analyses.^20^ ATTR-CM subtype (ATTRwt-CM or ATTRv-CM) was defined using ICD-10-CM coding algorithms. To be considered as having ATTRv-CM, a patient was required to have ≥2 heredofamilial amyloidosis codes (on separate days) during the identification period. If a patient had ATTRwt-CM codes and <1 heredofamilial amyloidosis codes during the identification period, the patient was considered to have ATTRwt-CM. Baseline clinical variables assessed at or within 90 days of ATTR-CM diagnosis included chronic kidney disease, diabetes mellitus, hypertension, coronary artery disease, obesity, stroke, systolic heart failure (HF), and diastolic HF. Diastolic HF was included as a covariate to more accurately estimate the impact of sex and race on the target outcomes. The need for a loop diuretic was approximated by the loop diuretic (furosemide, torsemide, bumetanide) dose at baseline, categorized as none (no loop diuretic use), minor or mild (furosemide <59 mg; torsemide <29 mg; bumetanide <1.4 mg), or moderate or severe (furosemide ≥60 mg; torsemide ≥30 mg; bumetanide ≥1.5 mg total daily dose).

### STATISTICAL ANALYSIS

Baseline characteristics were summarized by sex and race. The Kaplan-Meier method, stratified by sex and race, was used to estimate cumulative incidence of tafamidis initiation at 3, 6, 9, and 12 months post-diagnosis, as well as to estimate event-free survival for CVH or all-cause death. Multivariable Cox proportional hazards models were fit to model time to tafamidis initiation and, separately, time to CVH or all-cause death. White men served as the reference population in both Cox proportional hazards models. Covariates included sex, race, SDI, ATTR-CM type, year of diagnosis, age at diagnosis, US region, insurance type, need for a loop diuretic, and select comorbidities (chronic kidney disease, chronic obstructive pulmonary disease, diabetes mellitus, diastolic HF, hypertension, obesity, stroke, systolic HF). Prespecified interaction terms were sex × race, sex × SDI, and race × ATTR-CM type. Pairwise differences by race and sex (both overall and stratified by ATTR-CM type and SDI) were estimated using least square means derived from these models, which estimate adjusted survival outcomes by averaging over all other model covariates. Comparisons are presented using hazard ratios (HRs) and nominal *P* values (*P*<0.001 was considered significant to account for multiple pairwise comparisons across sex, race, ATTR-CM subtype, and SDI strata).

## RESULTS

### PATIENTS

A total of 59,330 patients with amyloid codes were identified during the study period; 11,311 met eligibility criteria after exclusions (**Supplemental Figure 1**), including a priori exclusion of 953 patients with AL amyloidosis alone, 653 with MM alone, and 1,851 with both. Overall, 63.9% of patients were male, 52.0% were White, 29.6% were Black, 18.4% were other/unknown races, and 91.3% had ATTRwt-CM. Mean (SD) age at diagnosis was 73.5 (11.6) years for men and 72.4 (13.1) years for women. Only White and Black patients (n=9,223) were included in the remaining analyses due to the small sample size and heterogeneity of other groups. In this population, 64.9% of patients were male, 63.7% were White, and 36.3% were Black (**Table 1**). Black patients were younger (mean [SD], 71.1 [12.6] years vs 76.1 [10.3] years), had higher rates of ATTRv-CM (12.6% vs 6.0%), had a higher degree of social deprivation (SDI 81-100, 39.7% vs 12.5%), and had a higher rate of moderate-to-severe loop diuretic use (furosemide ≥60 mg; 19.2% vs 12.3%) than White patients, respectively. In addition, more Black patients lived in the South compared with White patients (33.7% vs 17.4%, respectively). Substantially greater proportions of Black patients had chronic kidney disease (58.9% vs 41.5%), diabetes mellitus (52.4% vs 33.3%), and systolic HF (47.5% vs 33.0%) compared with White patients, respectively. Women (n=3,241) had a higher degree of social deprivation (SDI 81-100, 26.1% vs 20.3%) and greater proportions had stroke (37.4% vs 26.2%), while smaller proportions had COPD (50.5% vs 62.0%) and systolic HF (31.6% vs 41.9%) compared with men (n=5,982).

**Table 1.** Baseline Demographics and Patient Characteristics by Sex and Race^a^.

|  | Overall |  | Men |  | Women |  |
| --- | --- | --- | --- | --- | --- | --- |
| Characteristic | White<br>(n=5,877) | Black<br>(n=3,346) | White<br>(n=4,125) | Black<br>(n=1,857) | White<br>(n=1,752) | Black<br>(n=1,489) |
| **Age at diagnosis** |  |  |  |  |  |  |
| Mean (SD), years | 76.1 (10.3) | 71.1 (12.6) | 76.5 (9.6) | 70.3 (12.2) | 75.1 (11.6) | 72.2 (13.0) |
| **ATTR-CM type, n (%)** |  |  |  |  |  |  |
| ATTRwt-CM | 5,527 (94.0) | 2,923 (87.4) | 3,899 (94.5) | 1,614 (86.9) | 1,628 (92.9) | 1,309 (87.9) |
| ATTRv-CM | 350 (6.0) | 423 (12.6) | 226 (5.5) | 243 (13.1) | 124 (7.1) | 180 (12.1) |
| **US region, n (%)** |  |  |  |  |  |  |
| Northeast | 2,519 (42.9) | 1,094 (32.7) | 1,843 (44.7) | 558 (30.1) | 676 (38.6) | 536 (36.0) |
| Midwest | 1,455 (24.8) | 925 (27.6) | 1,019 (24.7) | 545 (29.4) | 436 (24.9) | 380 (25.5) |
| South | 1,022 (17.4) | 1,129 (33.7) | 665 (16.1) | 644 (34.7) | 357 (20.4) | 485 (32.6) |
| West | 878 (14.9) | 197 (5.9) | 596 (14.5) | 109 (5.9) | 282 (16.1) | 88 (5.9) |
| **Year of diagnosis, n (%)<sup>b</sup>** |  |  |  |  |  |  |
| 2019 | 626 (10.7) | 361 (10.8) | 435 (10.6) | 194 (10.5) | 191 (10.9) | 167 (11.2) |
| 2020 | 925 (15.7) | 552 (16.5) | 621 (15.1) | 303 (16.3) | 304 (17.4) | 249 (16.7) |
| 2021 | 1,187 (20.2) | 730 (21.8) | 833 (20.2) | 401 (21.6) | 354 (20.2) | 329 (22.1) |
| 2022 | 1,305 (22.2) | 813 (24.3) | 910 (22.1) | 464 (24.5) | 395 (22.6) | 349 (23.4) |
| 2023 | 1,394 (23.7) | 711 (21.3) | 1,011 (24.5) | 396 (21.3) | 383 (21.9) | 315 (21.2) |
| 2024 | 440 (7.5) | 179 (5.4) | 315 (7.6) | 99 (5.3) | 125 (7.1) | 80 (5.4) |
| **Medical insurance type, n (%)** |  |  |  |  |  |  |
| Medicare | 4,925 (83.8) | 2,467 (73.7) | 3,473 (84.2) | 1,336 (71.9) | 1,452 (82.9) | 1,131 (76.0) |
| Commercial | 658 (11.2) | 408 (12.2) | 482 (11.7) | 273 (14.7) | 176 (10.1) | 135 (9.1) |
| Medicaid | 289 (4.9) | 470 (14.1) | 166 (4.0) | 247 (13.3) | 123 (7.0) | 223 (15.0) |
| Unknown | 5 (<0.1) | 1 (<0.1) | 4 (0.1) | 1 (<0.1) | 1 (<0.1) | 0 |
| **SDI, n (%)** |  |  |  |  |  |  |
| ≤80 | 5,105 (86.9) | 2,012 (60.1) | 3,606 (87.4) | 1,134 (61.1) | 1,499 (85.6) | 878 (59.0) |
| >80 | 732 (12.5) | 1,327 (39.7) | 495 (12.1) | 719 (38.8) | 237 (13.7) | 608 (40.9) |
| Unknown | 40 (0.7) | 7 (0.2) | 24 (0.6) | 4 (0.2) | 16 (0.9) | 3 (0.2) |
| **Total daily equivalent oral furosemide use, n (%)** |  |  |  |  |  |  |
| No loop diuretic use | 2,875 (48.9) | 1,484 (44.4) | 1,886 (45.7) | 807 (43.5) | 989 (56.5) | 677 (45.5) |
| Furosemide <59 mg | 2,280 (38.8) | 1,219 (36.4) | 1,673 (40.6) | 665 (35.8) | 607 (34.6) | 554 (37.2) |
| Furosemide ≥60 mg | 722 (12.3) | 643 (19.2) | 566 (13.7) | 385 (20.7) | 156 (8.9) | 258 (17.3) |
| **Comorbidity, n (%)** |  |  |  |  |  |  |
| CKD | 2,437 (41.5) | 1,971 (58.9) | 1,774 (43.0) | 1,128 (60.7) | 663 (37.8) | 843 (56.6) |
| COPD | 3,478 (59.2) | 1,867 (55.8) | 2,631 (63.8) | 1,078 (58.1) | 847 (48.3) | 789 (52.3) |
| Diabetes mellitus | 1,958 (33.3) | 1,752 (52.4) | 1,379 (33.4) | 962 (51.8) | 579 (33.1) | 790 (53.1) |
| Diastolic heart failure | 2,913 (49.6) | 1,690 (50.5) | 2,024 (49.1) | 871 (46.9) | 889 (50.7) | 819 (55.0) |
| Hypertension | 4,984 (84.8) | 3,094 (92.5) | 3,486 (84.5) | 1,705 (91.8) | 1,498 (85.5) | 1,389 (93.3) |
| Obesity | 1,423 (24.2) | 1,102 (32.9) | 938 (22.7) | 546 (29.4) | 485 (27.7) | 556 (37.3) |
| Stroke | 1,692 (28.8) | 1,087 (32.5) | 1,031 (25.0) | 537 (28.9) | 661 (37.7) | 550 (36.9) |
| Systolic heart failure | 1,941 (33.0) | 1,589 (47.5) | 1,514 (36.7) | 991 (53.4) | 427 (24.4) | 598 (40.2) |

<sup>a</sup>Only White and Black patients were included in this analysis; patients of other races were excluded (n=2088).
<sup>b</sup>The patient identification period was from May 3, 2019, to June 30, 2024.
ATTR-CM = transthyretin amyloid cardiomyopathy; ATTRv-CM = variant transthyretin amyloid cardiomyopathy; ATTRwt-CM = wild-type transthyretin amyloid cardiomyopathy; CKD = chronic kidney disease; COPD = chronic obstructive pulmonary disease; SD = standard deviation; SDI = Social Deprivation Index.

### TIME TO TAFAMIDIS INITIATION

Women had a significantly lower cumulative incidence of tafamidis initiation compared with men (*P*<0.001). Within 3 months of diagnosis, the cumulative incidence of tafamidis initiation was 9.9% among women and 19.9% among men. This gap persisted at 12 months, with the cumulative incidence of tafamidis initiation among women and men at 14.5% and 28.5%, respectively. White patients had a higher cumulative incidence of tafamidis initiation within 3 months of diagnosis compared with Black patients (18.3% vs 16.0%). There was narrowing of this gap by 12 months (25.1% vs 24.6%); however, when stratified by both race and sex, White men had the highest cumulative incidence of tafamidis initiation at 12 months (31.0%), followed by Black men (26.7%), Black women (22.0%), and White women (11.4%) (**Figure 1**).

**Figure 1.**
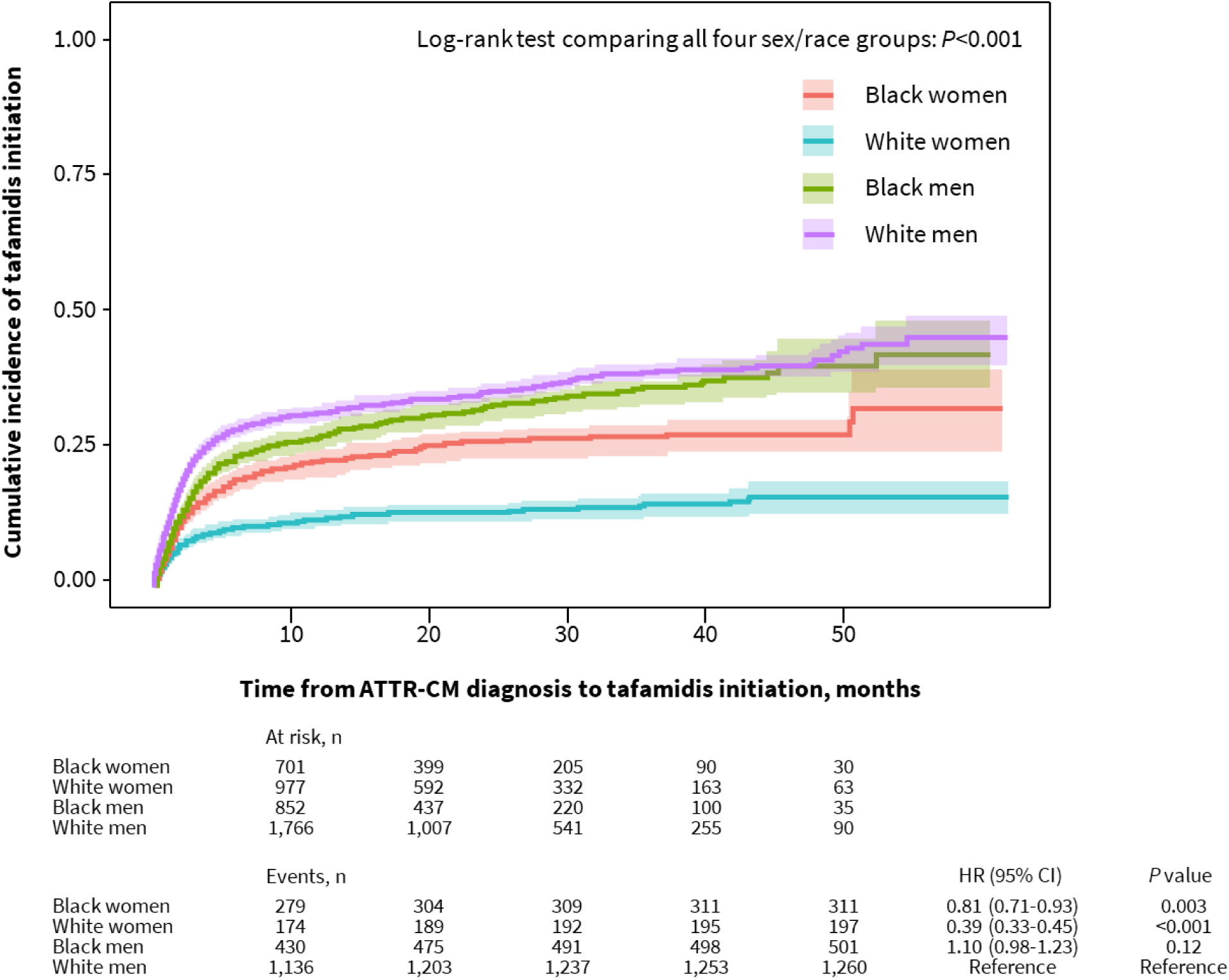
Cumulative Incidence of Tafamidis Initiation by Sex and Race. Diagnosis was defined as the earliest occurrence of the 2 ATTR-CM diagnosis codes, and the treatment index date was defined as the first tafamidis claim. The table shows the number of patients at risk (not yet on treatment) and cumulative events (treatment initiations) at 0, 10, 20, 30, 40, and 50 months. ATTR-CM = transthyretin amyloid cardiomyopathy; HR = hazard ratio.

Results from the Cox model of time to tafamidis initiation are presented in **Supplemental Table 2**. Overall, pairwise contrasts of time to tafamidis initiation by race and sex demonstrated significant disparities, with White women having the longest time to tafamidis initiation after adjusting for covariates (*P*<0.001) (**Supplemental Table 3**). White women initiated tafamidis later than White men (HR: 0.39 [95% CI: 0.33-0.45]; *P*<0.001) as well as both Black men (HR: 2.83 [95% CI: 2.38-3.37]; *P*<0.001) and Black women (HR: 2.10 [95% CI: 1.73-2.54]; *P*<0.001). Black women initiated tafamidis later than both Black men (HR: 0.74 [95% CI: 0.64-0.86]; *P*<0.001) and White men (HR: 0.81 [95% CI: 0.71-0.93]; *P*=0.003). These patterns were largely consistent across ATTR-CM subtypes, with notable exceptions: Black men initiated tafamidis earlier than White men (HR: 1.74 [95% CI: 1.32-2.30]; *P*<0.001) only among those with ATTRv-CM, and Black women initiated tafamidis later than White men (HR: 0.78 [95% CI: 0.68-0.90]; *P*<0.001) only among those with ATTRwt-CM.

Across SDI strata for low (SDI ≤80) and high social vulnerability (SDI >80), White women initiated tafamidis later than White men, while both Black men and Black women initiated tafamidis earlier compared with White women (**Supplemental Table 3**).

### TIME TO CVH OR DEATH

Kaplan-Meier curves demonstrated significant differences in time to CVH or death stratified by race and sex (*P*<0.001) (**Figure 2**). Event-free survival at 12 months was lowest for Black women (42.9%), followed by Black men (46.8%), White women (48.6%), and White men (54.4%). Accordingly, Black women had the shortest median (95% CI) time to CVH or death (8.0 months [6.8-10.0]), followed by Black men (9.9 months [8.8-12.0]), White women (11.0 months [9.6-13.0]), and White men (15.0 months [14.0-16.0]) (**Table 2**).

**Figure 2.**
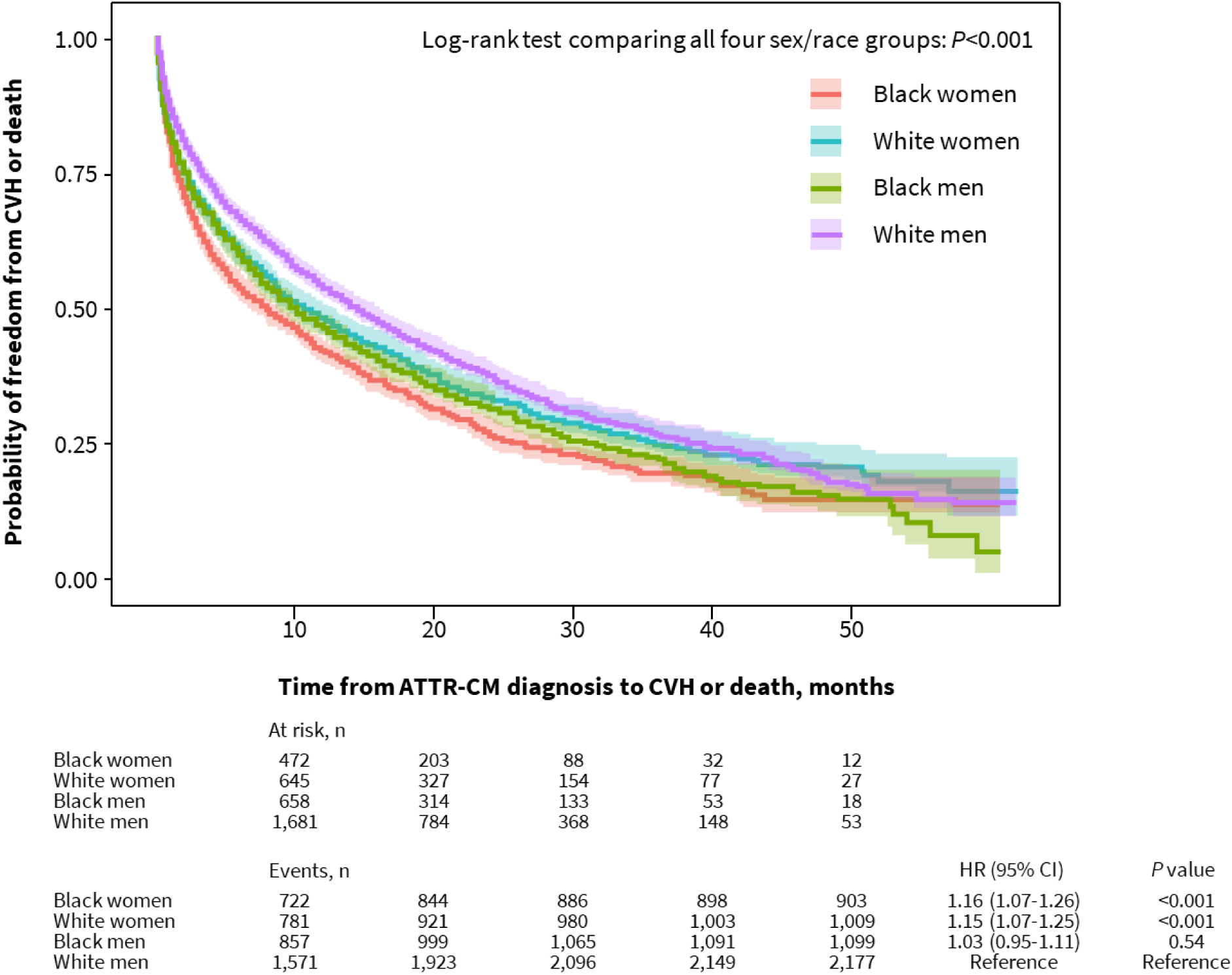
Time to CVH or Death by Sex and Race. Diagnosis was defined as the earliest occurrence of the 2 ATTR-CM diagnosis codes, CVH was defined as inpatient encounters with a cardiovascular-related ICD-10-CM code, and death was identified using claims-based mortality flags. The table shows the number of patients at risk (who have not yet experienced CVH or death) and cumulative events (CVH or death) at 0, 10, 20, 30, 40, and 50 months. ATTR-CM = transthyretin amyloid cardiomyopathy; CVH = cardiovascular-related hospitalization; HR = hazard ratio; ICD-10-CM = International Classification of Diseases, Tenth Revision, Clinical Modification.

**Table 2.**
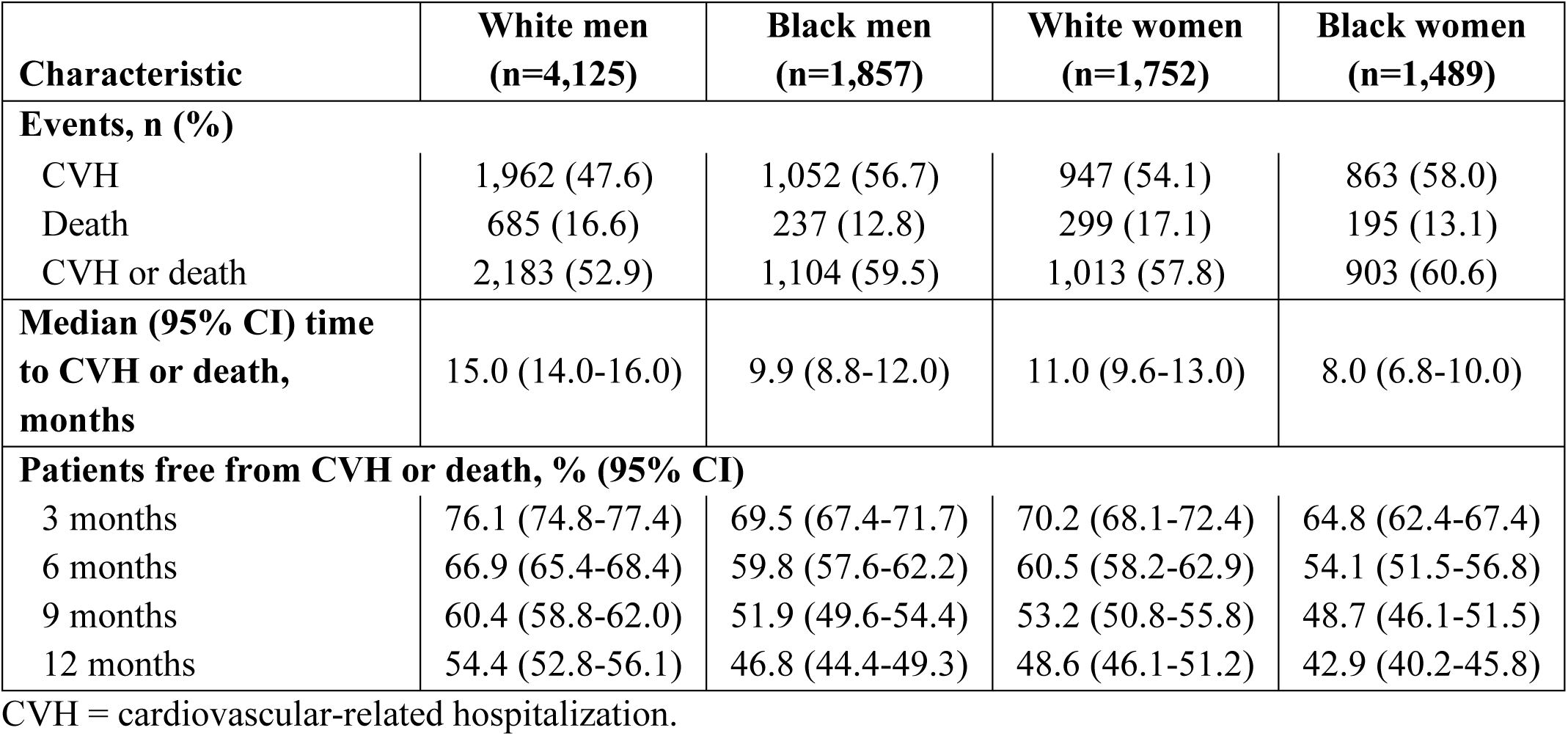
CVH or Death by Sex and Race.

| **Characteristic** | **White men**<br>**(n=4,125)** | **Black men**<br>**(n=1,857)** | **White women**<br>**(n=1,752)** | **Black women**<br>**(n=1,489)** |
| --- | --- | --- | --- | --- |
| **Events, n (%)** |  |  |  |  |
| CVH | 1,962 (47.6) | 1,052 (56.7) | 947 (54.1) | 863 (58.0) |
| Death | 685 (16.6) | 237 (12.8) | 299 (17.1) | 195 (13.1) |
| CVH or death | 2,183 (52.9) | 1,104 (59.5) | 1,013 (57.8) | 903 (60.6) |
| **Median (95% CI) time**<br>**to CVH or death,**<br>**months** | 15.0 (14.0-16.0) | 9.9 (8.8-12.0) | 11.0 (9.6-13.0) | 8.0 (6.8-10.0) |
| **Patients free from CVH or death, % (95% CI)** |  |  |  |  |
| 3 months | 76.1 (74.8-77.4) | 69.5 (67.4-71.7) | 70.2 (68.1-72.4) | 64.8 (62.4-67.4) |
| 6 months | 66.9 (65.4-68.4) | 59.8 (57.6-62.2) | 60.5 (58.2-62.9) | 54.1 (51.5-56.8) |
| 9 months | 60.4 (58.8-62.0) | 51.9 (49.6-54.4) | 53.2 (50.8-55.8) | 48.7 (46.1-51.5) |
| 12 months | 54.4 (52.8-56.1) | 46.8 (44.4-49.3) | 48.6 (46.1-51.2) | 42.9 (40.2-45.8) |

CVH = cardiovascular-related hospitalization.

Results from the Cox model of time to CVH or death are presented in **Supplemental Table 4**. Overall, pairwise contrasts of race and sex for time to CVH or death demonstrated that both White women (HR: 1.15 [95% CI: 1.07-1.25]; *P*<0.001) and Black women (HR: 1.16 [95% CI: 1.07-1.26]; *P*<0.001) had a significantly greater risk of CVH or death compared with White men (**Supplemental Table 5**). Patterns were consistent across ATTR-CM subtypes, with the exception of Black men with ATTRv-CM, who had a significantly lower risk of CVH or death compared with White women with ATTRv-CM (HR: 0.69 [95% CI: 0.56-0.86]; *P*<0.001). Among patients with low social vulnerability (SDI ≤80), sex disparities persisted, while there were no significant differences in patients with high social vulnerability (SDI >80) when compared by sex and race (**Supplemental Table 5**).

## DISCUSSION

In this national cohort of more than 11,000 patients with ATTR-CM, marked sociodemographic differences were observed in tafamidis initiation and subsequent clinical outcomes (**Central Illustration**). Our major findings include: 1) Women were significantly less likely than men to initiate tafamidis within 12 months of diagnosis, with White women having the lowest uptake. Black women had higher tafamidis initiation rates than White women, but remained disadvantaged compared with men. 2) Survival probabilities for CVH or death were lower among female patients and among Black patients. 3) There were notable differences in factors associated with time to tafamidis initiation compared with those associated with the clinical outcomes of CVH or death. During a time of increased disease awareness, less invasive diagnostic techniques, and available disease modifying therapies, inequities in the treatment and clinical outcomes of women and Black patients is a serious cause for concern.

**CENTRAL ILLUSTRATION.**
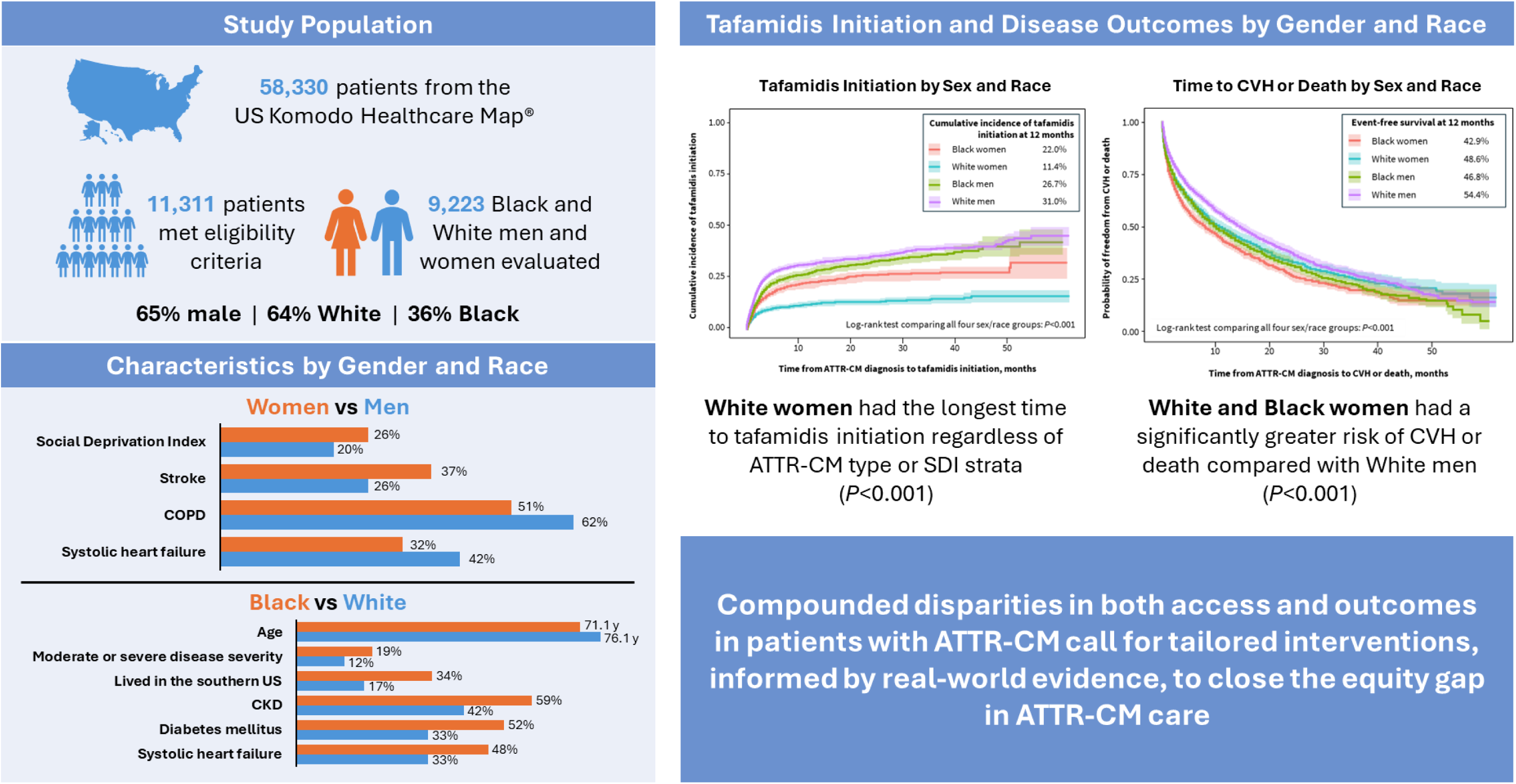
Disparities in Tafamidis Initiation and Outcomes. In this retrospective cohort analysis of 9,223 White and Black patients with ATTR-CM in the US, White women had the lowest cumulative incidence of tafamidis initiation within 12 months of diagnosis, followed by Black women, Black men, and White men. Black women had the lowest event-free survival at 12 months and shortest median time to CVH or death, followed by Black men, White women, and White men. Strategies to close the equity gap in ATTR-CM care are needed. ATTR-CM = transthyretin amyloid cardiomyopathy; CKD = chronic kidney disease; CVH = cardiovascular-related hospitalization; COPD = chronic obstructive pulmonary disease; SDI = Social Deprivation Index.

Approximately 36% of patients in the overall cohort were women. This is greater than the proportion of women observed in the Cardiac Amyloidosis Registry Study (CARS; 13%), but is more closely aligned with the proportion of women observed in the active ascertainment cohort of the Screening for Cardiac Amyloidosis with Nuclear Imaging in Minority Populations study (SCAN-MP; 31%), which had a recruitment target of 50% for women.^21-23^ The observation that White women had the lowest tafamidis initiation rates is particularly striking and suggests sex-related barriers independent of race. First, despite prospective studies of patients with heart failure showing closer to equal proportions of women and men with ATTR-CM by post- and ante-mortem analyses, ATTR-CM is wrongly considered a predominantly male disease, and there is a lack of real-world data on outcomes in female patients.^24,25^ Systematic underscreening, underdiagnosis, and underrepresentation in clinical trials have left the literature on sex differences in ATTR-CM limited and inconsistent, compounded by a lack of real-world data on outcomes in women. For example, some prior cohorts reported a higher age at ATTR-CM diagnosis in women compared with men,^17,18,26^ although this pattern was not observed in the present study. Similarly, reported differences in disease severity between sexes have been attenuated in studies accounting for body size.^17,18,27^

Our cohort offers an opportunity to address part of this data gap. Women have been underrepresented in ATTR-CM phase 3 clinical trials, accounting for <10% of enrolled patients,^6,28,29^ compared with 36% in the current cohort analysis. This higher representation may provide observations about female patients that may be more reflective of real-world clinical practice. The higher initiation of tafamidis among Black women compared with White women may reflect greater clinical vigilance due to awareness of the greater potential for Black individuals to be carriers of the p.Val142Ile variant, which is found almost exclusively in individuals of West African descent,^30-32^ though absolute rates remained lower than in men. Notably, 7.1% of White women in our cohort had ATTRv-CM. Together, these results underscore the need to address both structural and clinical drivers of unequal access.

Barriers to equitable quality of care and health outcomes, whether caused by affordability, technical literacy, or racism and bias, continue to pose a problem in the US.^33^ Despite the progressive increase in tafamidis initiation from 3 to 12 months post-diagnosis for all groups, women had higher risk of CVH or death at 12 months compared with White men. This disparity persisted after adjusting for tafamidis initiation, indicating that later-stage recognition, referral gaps, and limited access to advanced therapies may play important roles. More research is needed to identify additional mediators of the association between sex and clinical outcomes. The pivotal ATTR-ACT trial established tafamidis as the initial ATTR-CM disease modifying therapy which reduced mortality and hospitalizations,^6^ with the long-term extension studies showing sustained survival benefit with early treatment initiation.^9^ Real-world studies have confirmed that delays in treatment initiation diminish survival benefits.^10^ Our results suggest that inequities in tafamidis access may translate into clinically meaningful survival gaps. The ATTR-CM subtype analysis revealed an important nuance regarding racial disparities in disease outcomes. Although Black patients with ATTRv-CM had lower adjusted risk of CVH or death compared with White patients with variant disease, raw survival analyses showed that Black women overall had the lowest 12-month event-free survival. This apparent discrepancy may reflect the influence of later-stage diagnosis and residual confounding not fully captured in claims-based adjustments (eg, physician experience, patient functional status, or disease severity). Future prospective studies incorporating imaging and biomarker staging will be required to clarify these differences and better understand how sex, race, and genetic subtype interact to shape outcomes.

Our findings expand on prior reports of delayed diagnosis and worse clinical outcomes in underrepresented minorities with ATTR-CM. A retrospective analysis of 231 patients with ATTR-CM demonstrated that Afro-Caribbean patients had more advanced disease at the time of diagnosis and experienced poor outcomes more frequently than White patients.^4^ The current analysis shows that Black patients had higher rates of comorbidities than White patients. While higher mortality rates have been reported in counties near amyloid centers, southern states, despite having a higher proportion of Black patients than other US regions, reported the lowest mortality rates. This is likely due to disparities in the diagnosis of ATTR-CM among Black patients.^15^ Earlier recognition of ATTR-CM in women and Black patients is critical, supported by guideline-directed diagnostic pathways.^34,35^ Clinical trials must prioritize inclusive enrollment to strengthen generalizability and reduce uncertainty that contributes to treatment inequity.^36,37^ Implementation studies are needed to evaluate strategies that close gender and racial gaps in tafamidis initiation and improve long-term outcomes.

Our analysis demonstrated a lack of SDI effect on tafamidis initiation, but there was an effect on clinical outcomes. Women with low social vulnerability had a greater risk of CVH or death compared with White men, while there was no difference in patients with high social vulnerability. This paradox may be partly explained by the use of patient assistance programs to acquire tafamidis, the status for which could not be discerned with the study data source. Eligibility criteria for patient assistance programs create a nonlinear relationship between income and treatment access: the lowest-income patients may obtain tafamidis through patient assistance programs, while middle-income patients—who exceed patient assistance program thresholds but cannot afford high out-of-pocket costs—face the greatest barriers. Prior research shows that 35% of patients receive tafamidis solely through the patient assistance program, with an additional 14% using both Medicare and the patient assistance program.^13^ Given that patients receiving tafamidis through the patient assistance program were not indicated in the Komodo database, we cannot determine the true treatment patterns across income levels in our cohort. This may explain why socioeconomic factors did not modify gender gaps while sex and race disparities remained prominent.

### LIMITATIONS

Several limitations merit consideration. Claims-based data are vulnerable to diagnostic misclassification and the results may underrepresent undocumented variables. Further, without a published validated ATTR-CM coding algorithm,^38^ additional limitations include the limited ability to discern ATTR-CM genotype, as the relevant codes are relatively new, not widely or consistently used, and not linked to follow-up genetic testing. Although patients with codes for AL amyloidosis or MM were excluded a priori, residual AL-CM contamination cannot be fully ruled out among patients coded with nonspecific amyloidosis codes (E85.4, E85.89), particularly those who did not receive disease-specific therapy. Disease severity could not be directly assessed; the loop diuretic dose was used as a proxy in the absence of imaging or biomarker data. There was also a lack of data on biomarkers, disease staging, and disease severity in the adjustments for the multivariable analyses. Our primary models estimated the direct disparity attributable to race and sex, adjusting for clinical phenotype variables—diastolic and systolic HF and loop diuretic dose—that typically reflect disease expression at or near the time of ATTR-CM diagnosis. This adjustment helps isolate disparities that are independent of measured need for a loop diuretic and phenotype, but may attenuate disparities that operate through later presentation, diagnostic coding, or clinical labeling that are themselves influenced by severity. Mortality ascertainment from claims may underestimate events, and the limitations of claims-based death capture should be considered when interpreting the all-cause death component of the composite endpoint. Residual confounding, including provider practice patterns and patient preferences, cannot be excluded. We could not discern patients who may have received tafamidis through the patient assistance program and as mentioned earlier, this may have impacted our estimates of the association between SDI and tafamidis use. Patient data were limited in parts of the Western US due to lack of Komodo Healthcare Map data captured in closed healthcare systems. Finally, multiple ad hoc comparisons can inflate the type I error.

## CONCLUSIONS

Sociodemographic factors significantly influenced tafamidis initiation and clinical outcomes in ATTR-CM. Women, particularly White women, were least likely to initiate tafamidis, while Black women had the poorest survival, underscoring compounded disparities in both access and outcomes. These findings, informed by real-world evidence, reveal inequities in access to care leading to poor clinical outcomes, and strategies need to be identified to close the equity gap in ATTR-CM care and patient outcomes. Addressing these inequities will require coordinated approaches spanning diagnosis, prescribing, payer policy, and research inclusivity, to ensure equitable delivery of disease-modifying treatment in ATTR-CM. Our findings may increase awareness of critical demographic disparities in ATTR-CM treatment and outcomes and identify opportunities to improve the equity in ATTR-CM treatment access, time to treatment initiation, and ultimately optimize clinical outcomes for all patients with ATTR-CM.

## ABBREVIATIONS AND ACRONYMS

ATTR-CM: transthyretin amyloid cardiomyopathy
ATTRv-CM: variant transthyretin amyloid cardiomyopathy
ATTRwt-CM: wild-type transthyretin amyloid cardiomyopathy
CVH: cardiovascular-related hospitalization
HF: heart failure
ICD-10-CM: International Classification of Diseases, Tenth Revision, Clinical Modification
SDI: Social Deprivation Index
TTR: transthyretin.

## DATA AVAILABILITY

The data that support the findings of this study were used under license from Komodo Health^®^ and derived from the Komodo Healthcare Map^®^. Due to data use agreements and its proprietary nature, restrictions apply regarding the availability of the data. Further information is available from the sponsor of the study, BridgeBio Pharma, Inc., San Francisco, CA, USA.

## CLINICAL PERSPECTIVES

- **COMPETENCY IN MEDICAL KNOWLEDGE:** In this large, real-world analysis of the effects of sex and race on tafamidis initiation and clinical outcomes in US patients with ATTR-CM, White and Black women had lowest cumulative incidence of treatment initiation, followed by Black and White men, and Black women had the lowest event-free survival at 12 months and shortest time to CVH or death, followed by Black men, White women, and White men.
- **TRANSLATIONAL OUTLOOK:** These real-world data reveal compounded disparities in both access to care and poor clinical outcomes for patients, particularly in women, with ATTR-CM. Strategies to close the equity gap in ATTR-CM care are needed.

## ACKNOWLEDGMENTS

This study was funded by BridgeBio, Pharma, Inc. (San Francisco, CA, USA). All authors contributed to and approved the manuscript. Writing and editorial assistance were provided by Anne Jacobson, MPharm, CMPP, of The Lockwood Group (Stamford, CT, USA), funded by BridgeBio Pharma, Inc.

## AUTHOR CONTRIBUTIONS

All the authors participated in the conception and design of the study and in the analysis and interpretation of data. All the authors participated in the preparation and final approval of the manuscript.

## DISCLOSURES

**NC-S:** Received honoraria from Alnylam Pharmaceuticals, BridgeBio Pharma, Inc., and Pfizer; **HG:** Received research grant support from Akcea Therapeutics (Ionis Pharmaceuticals/AstraZeneca), Alnylam Pharmaceuticals, BridgeBio Pharma, Inc., eMyosound, Pfizer, and Roche Diagnostics; consulting income from Alnylam Pharmaceuticals, AstraZeneca, BridgeBio Pharma, Inc., eMyosound, Merck, Novo Nordisk, Pfizer, and TD Cowen; stock option for Eko Health; research payments for clinical endpoint committees from Baim Institute for Clinical Research for Abbott, Beckman Coulter Diagnostics, Innolife, and Siemens, and from ACI/WCG Clinical for Abbott Laboratories and Alexion Pharmaceuticals; reimbursement/honoraria from Inova Schar Heart and Vascular; and support from Women As One for the 2025 Women As One Mentorship Awards; **AMR** and **LH:** Employees and stockholders of BridgeBio Pharma, Inc.; **MU:** Employee and stockholder of BridgeBio Pharma, Inc., and stockholder of Pfizer; **BZ, XG, EN,** and **AK:** Contractors for BridgeBio Pharma, Inc.; **MKD:** Received honoraria from Alnylam Pharmaceuticals, AstraZeneca, BridgeBio Pharma, Inc., Novo Nordisk, and Pfizer; and research grant support from Pfizer.

## SUPPLEMENTAL APPENDIX

**Supplemental Table 1.**
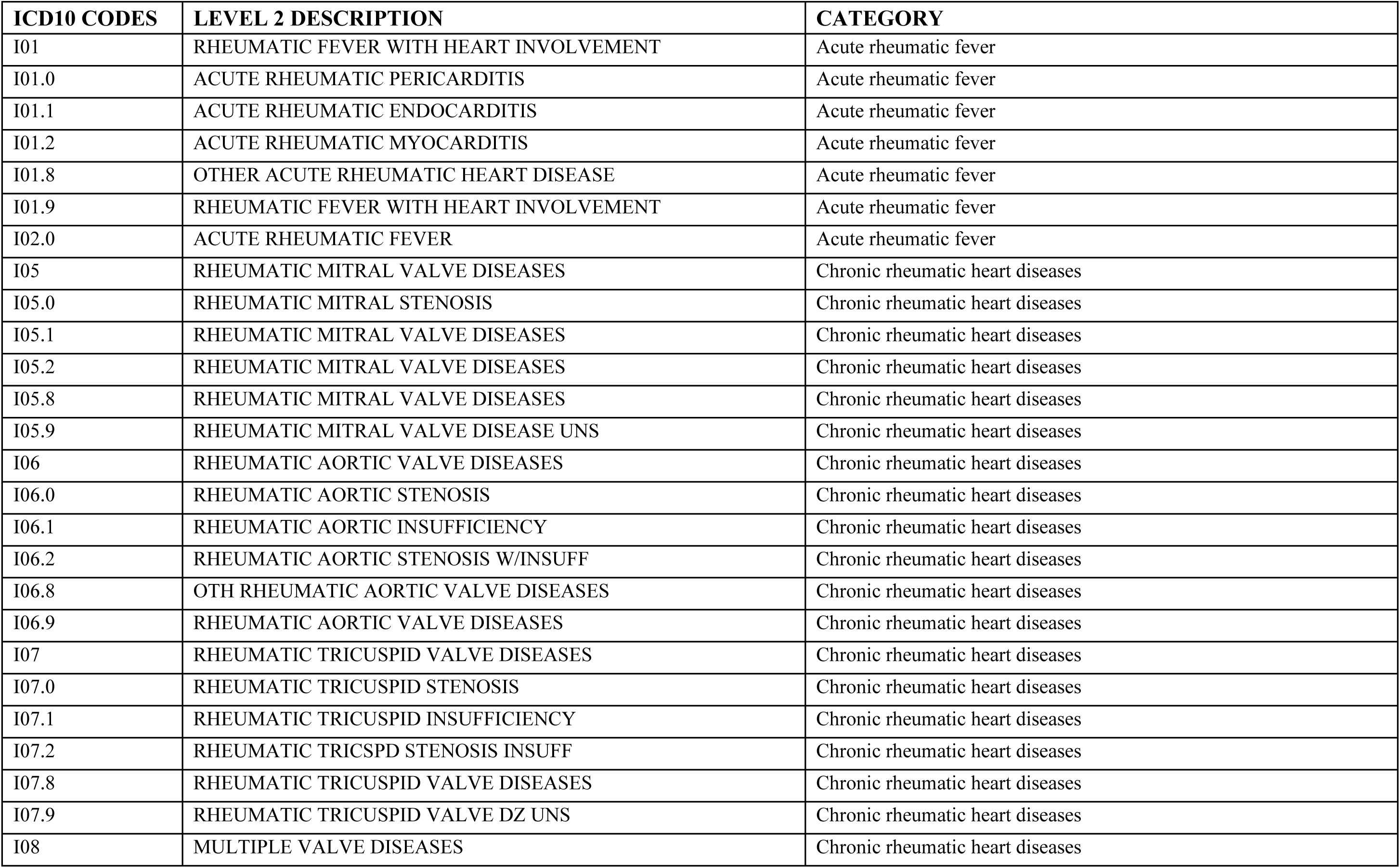
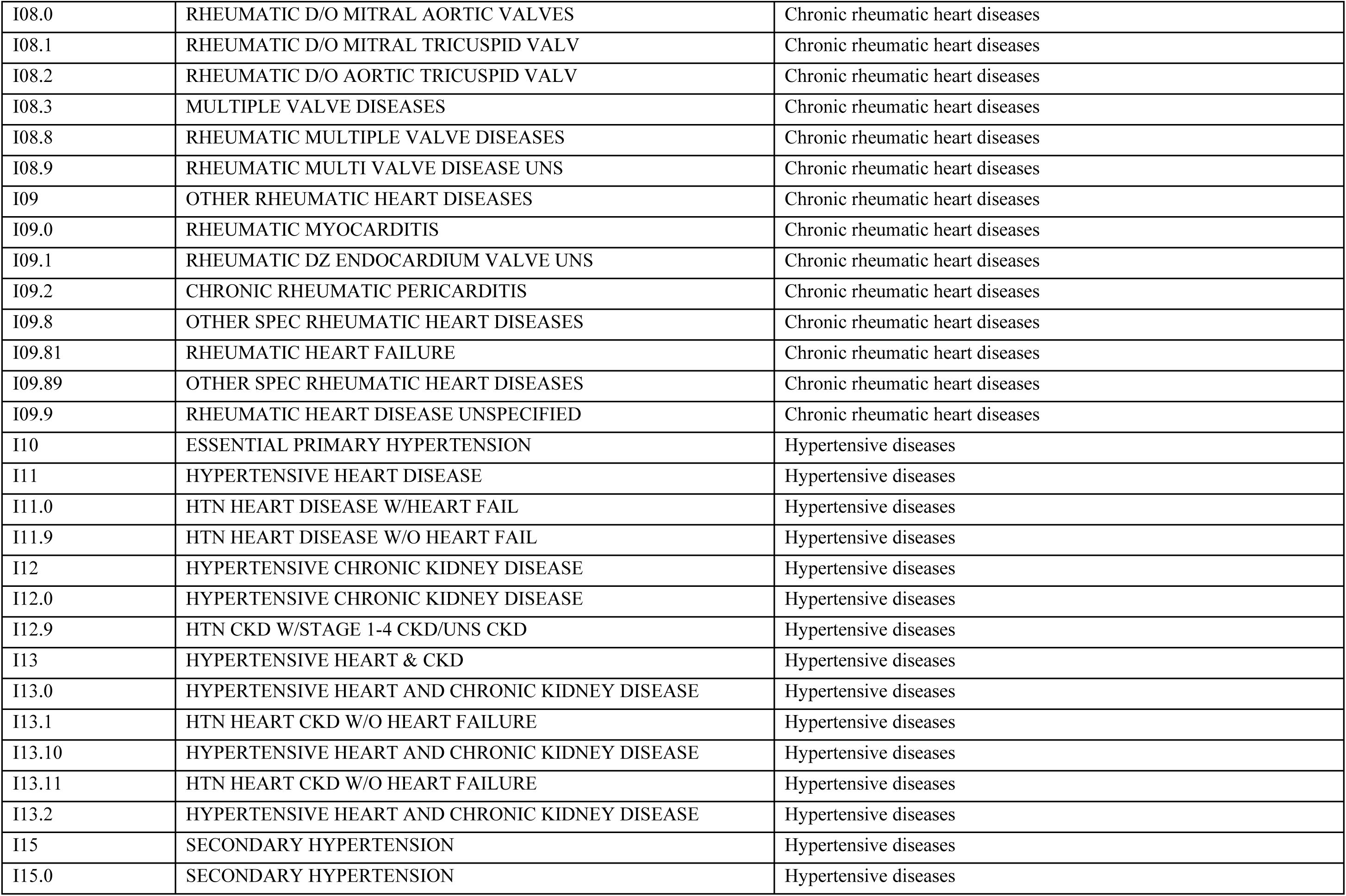
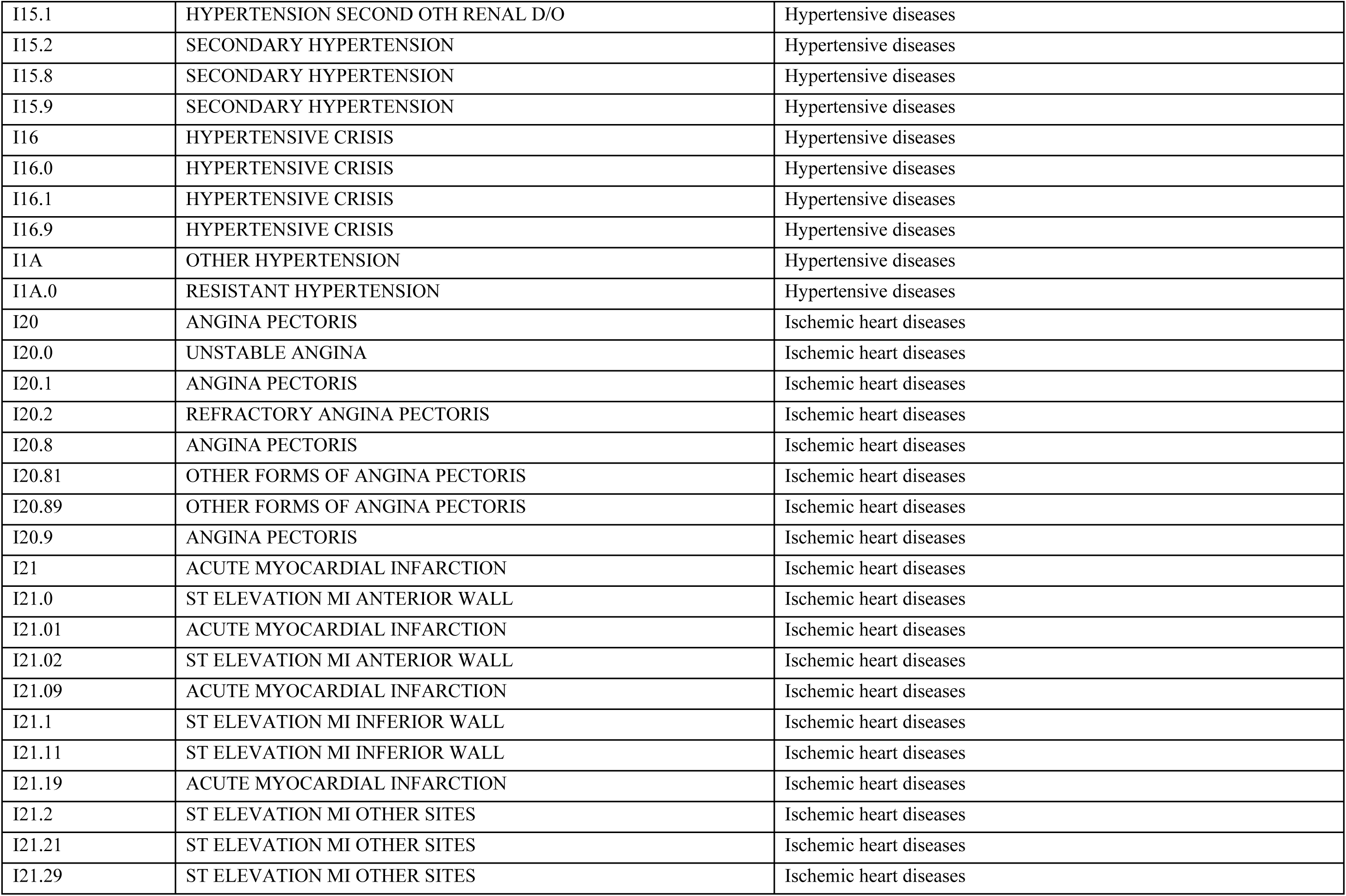
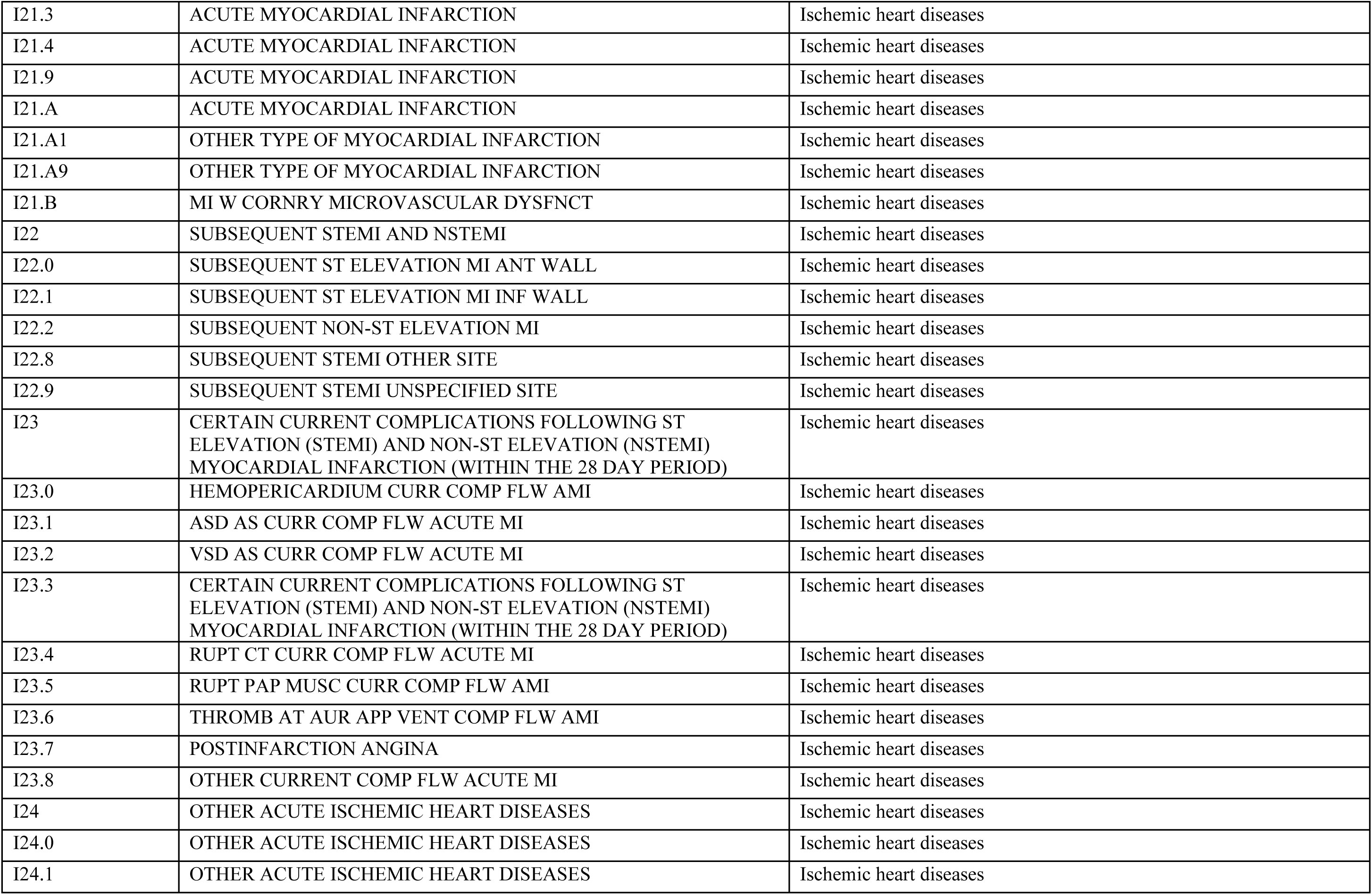
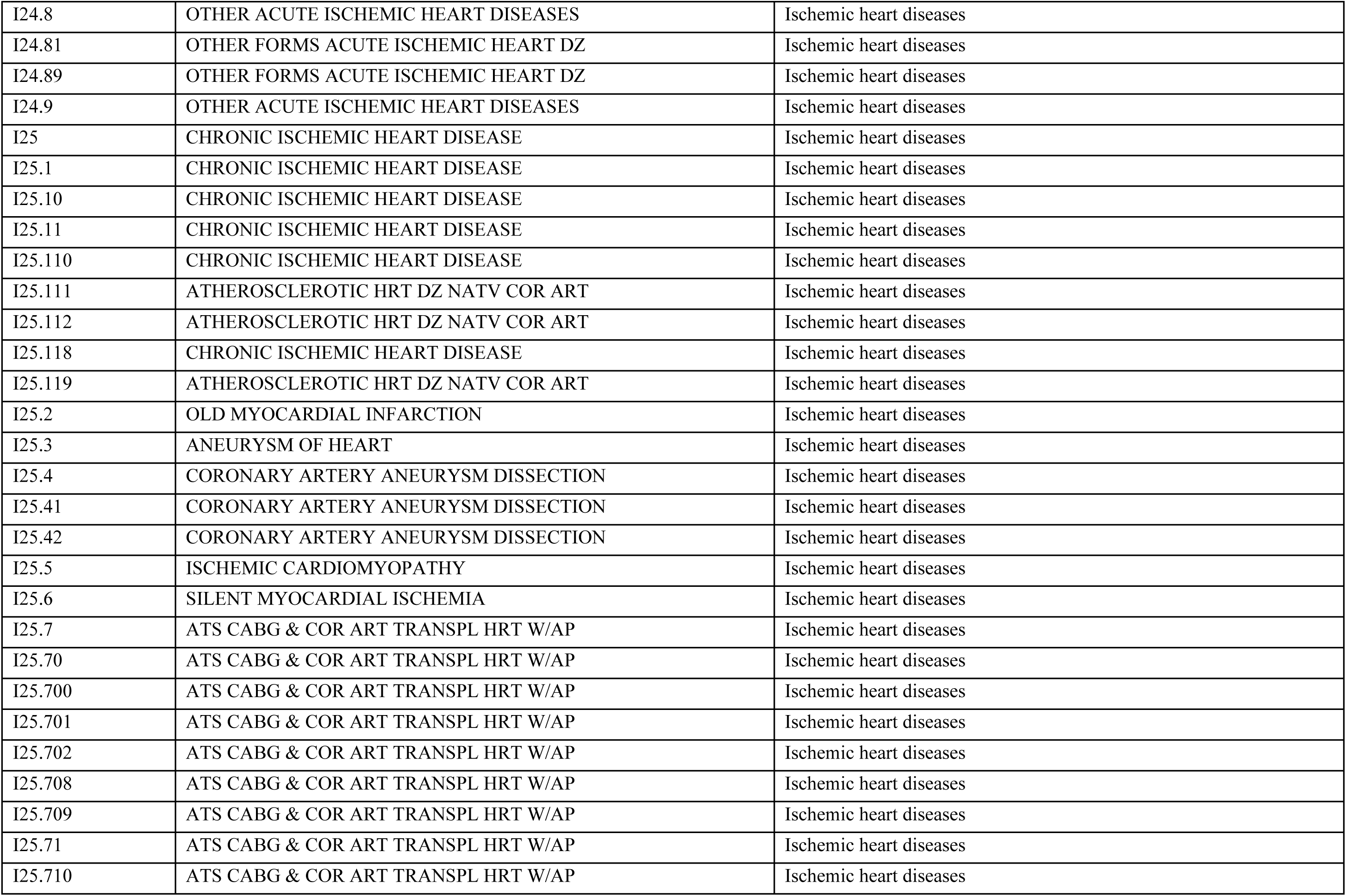
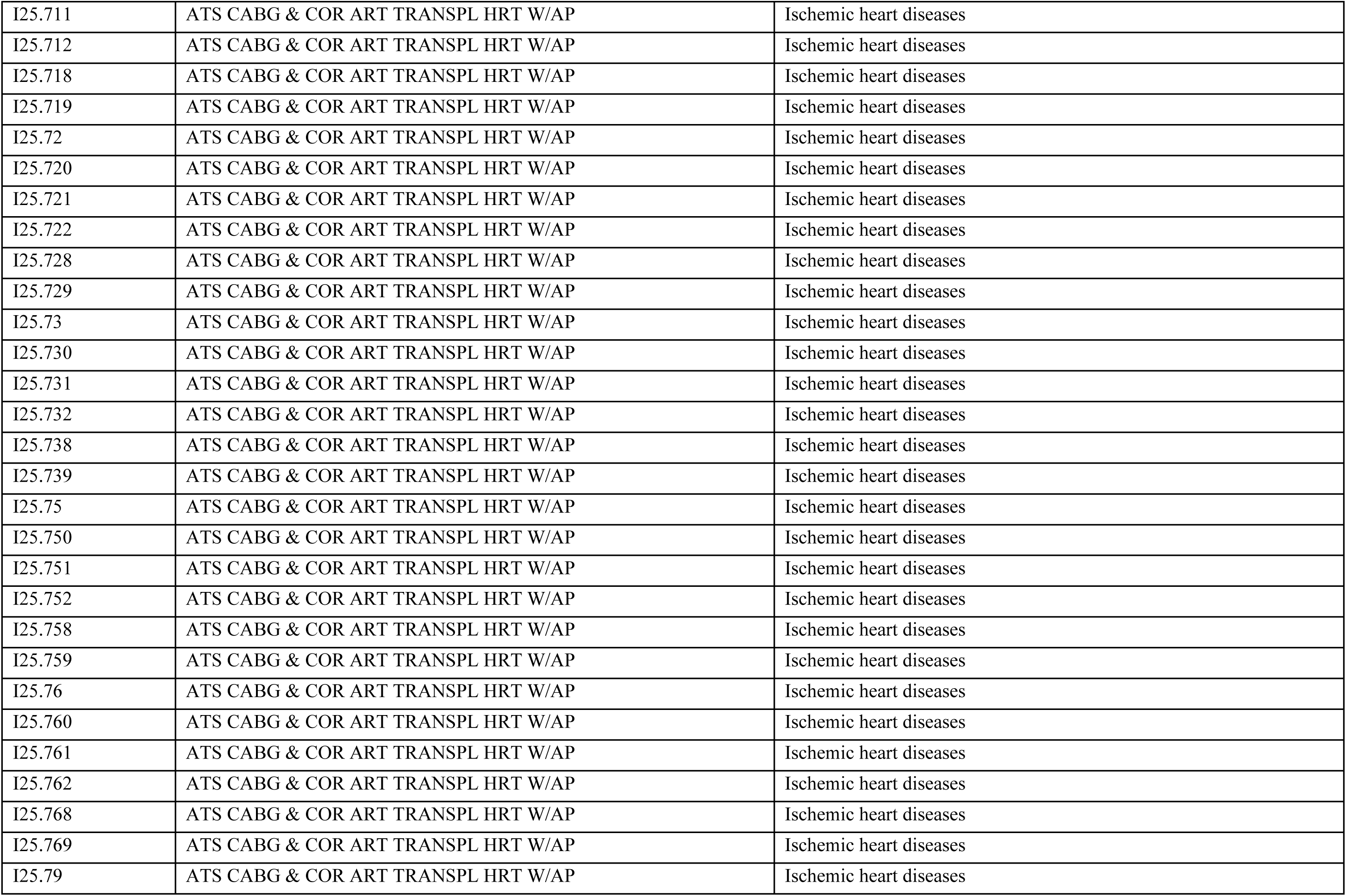
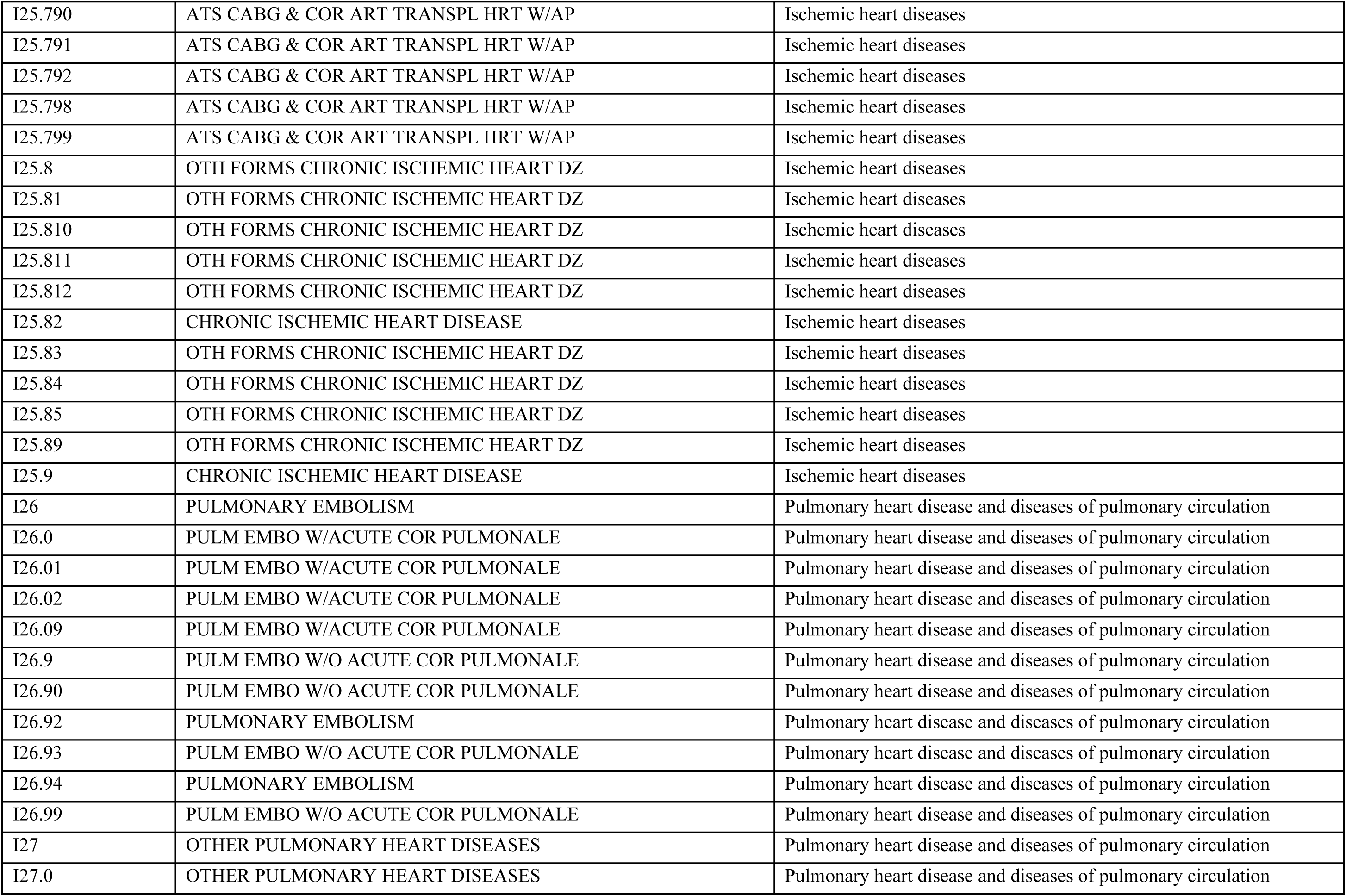
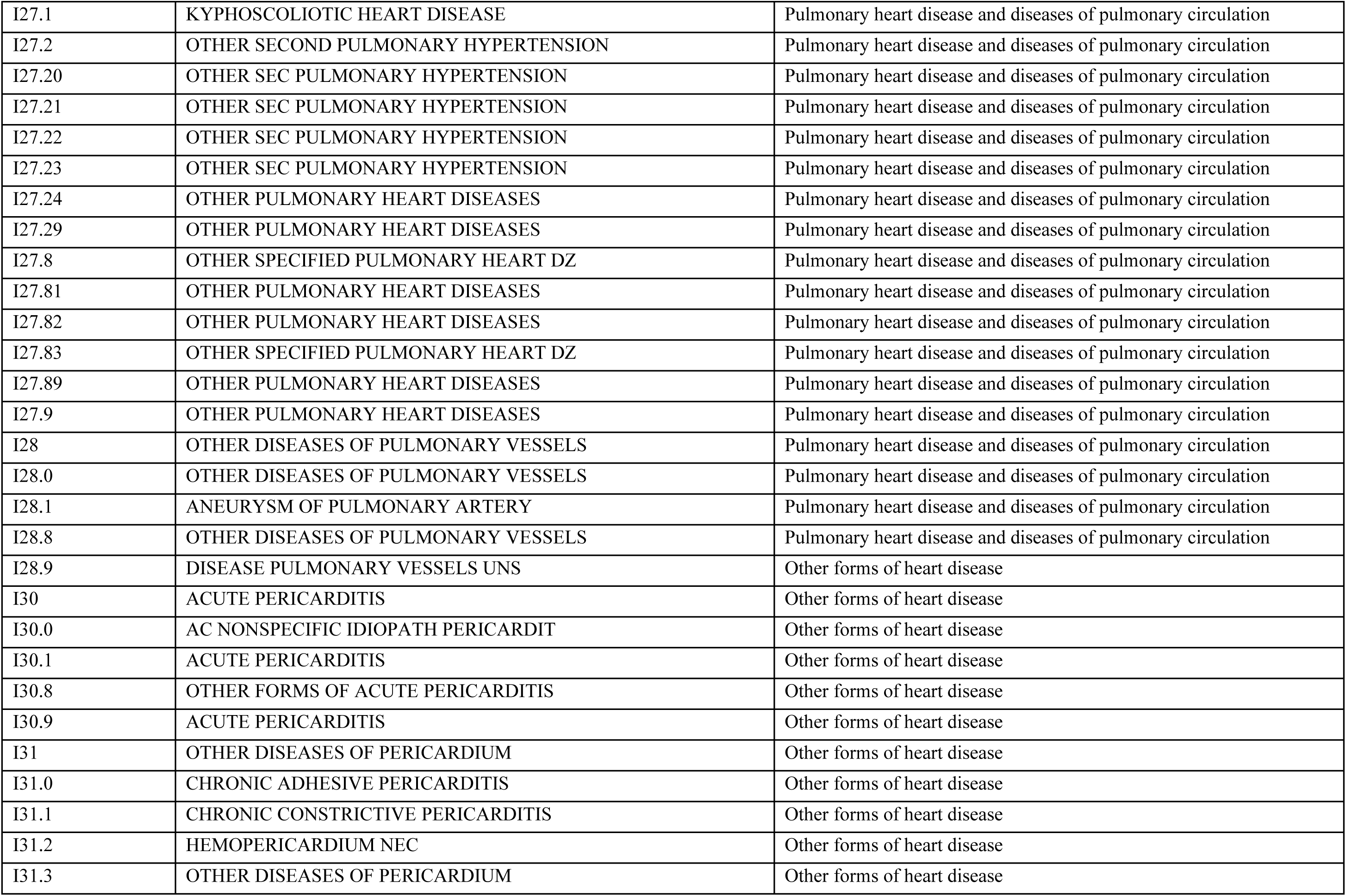
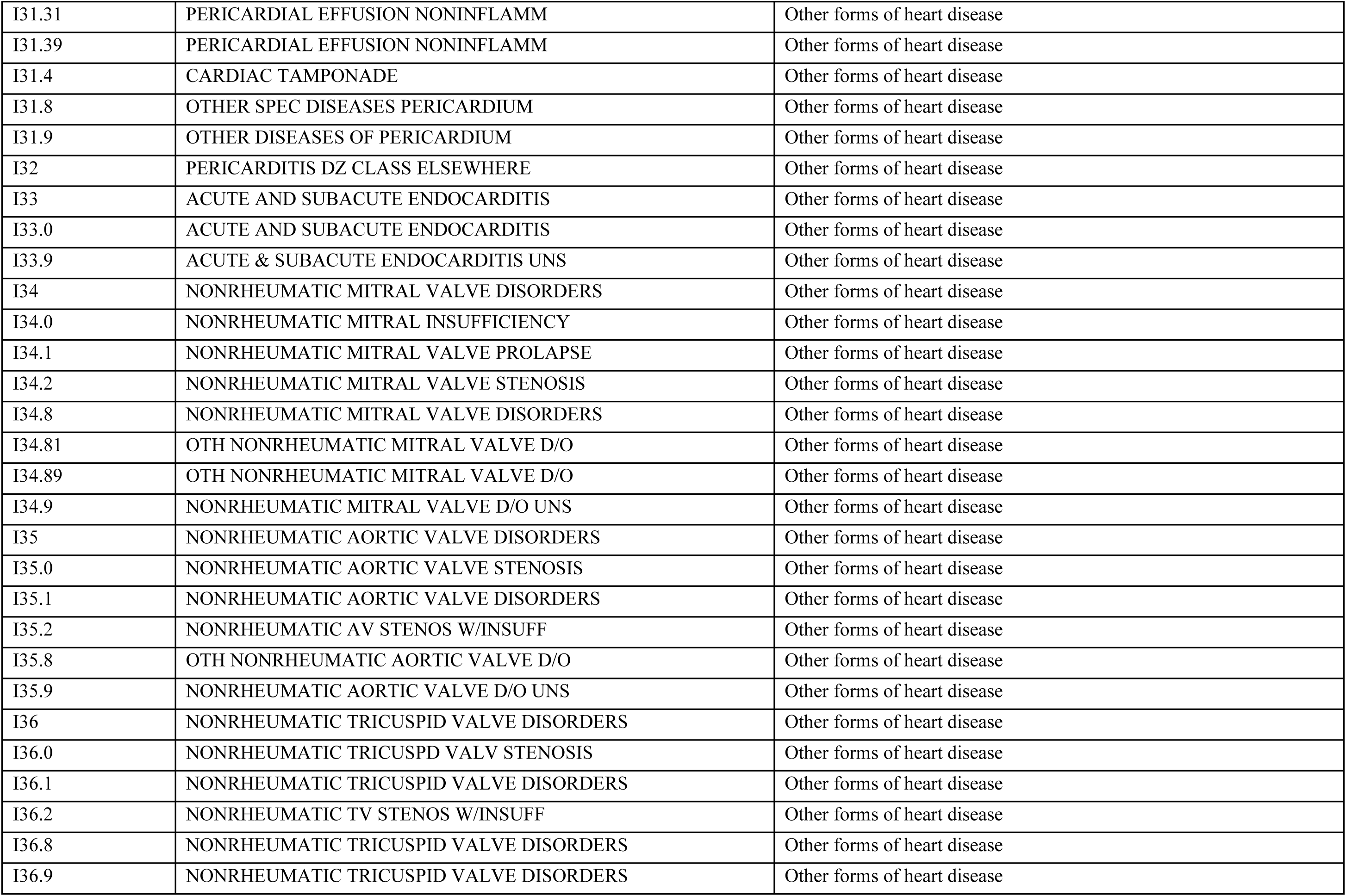
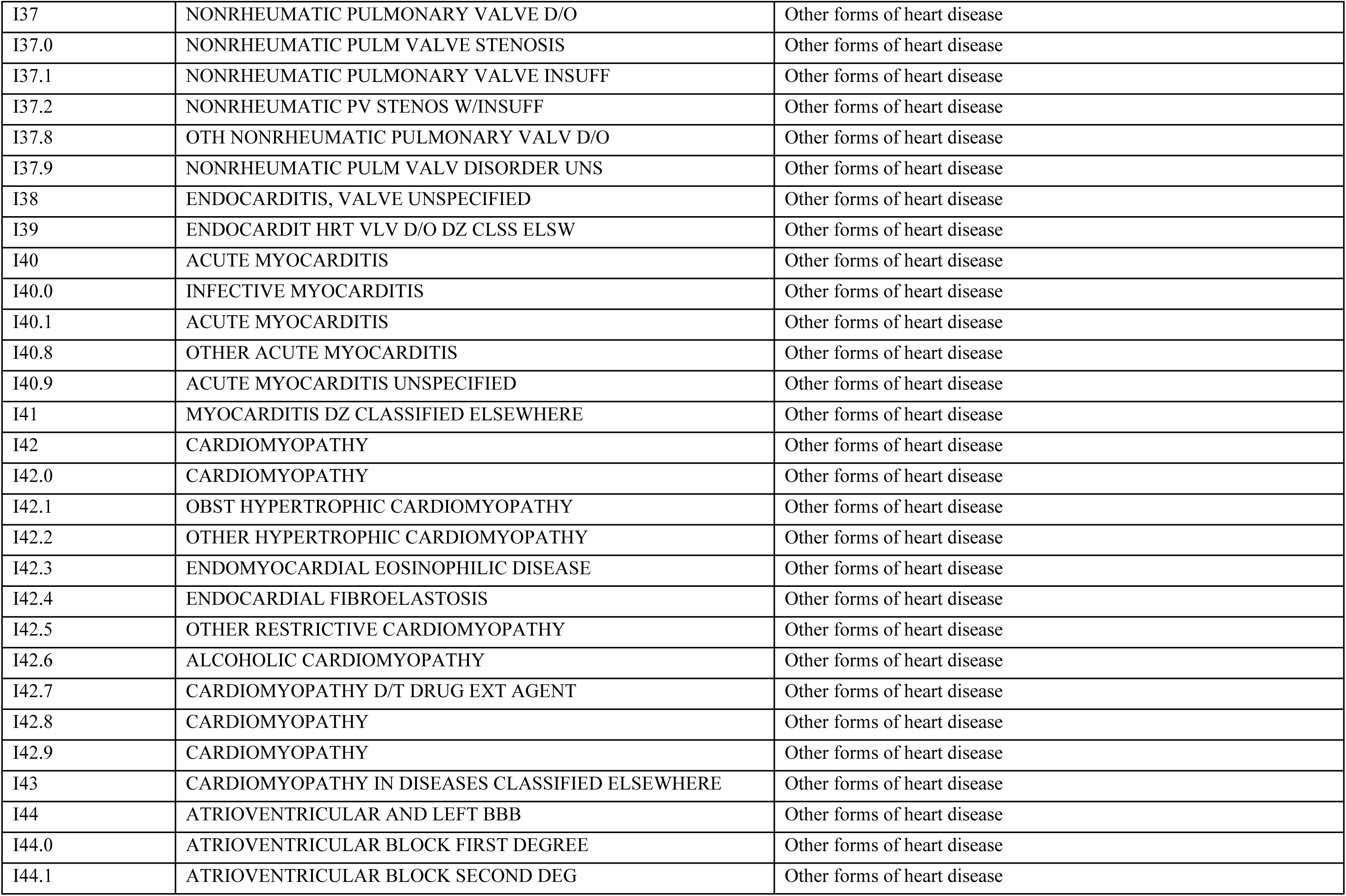
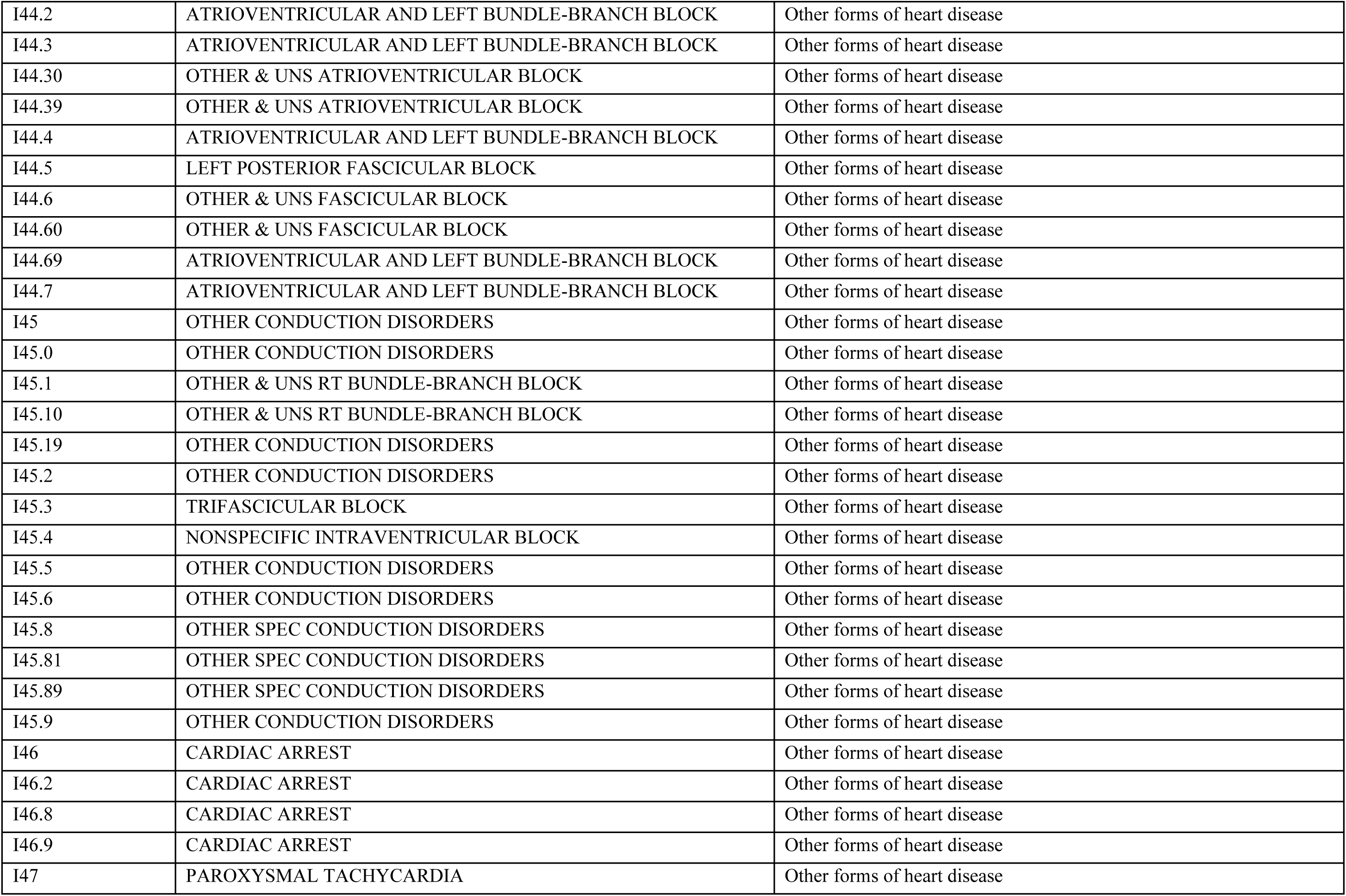
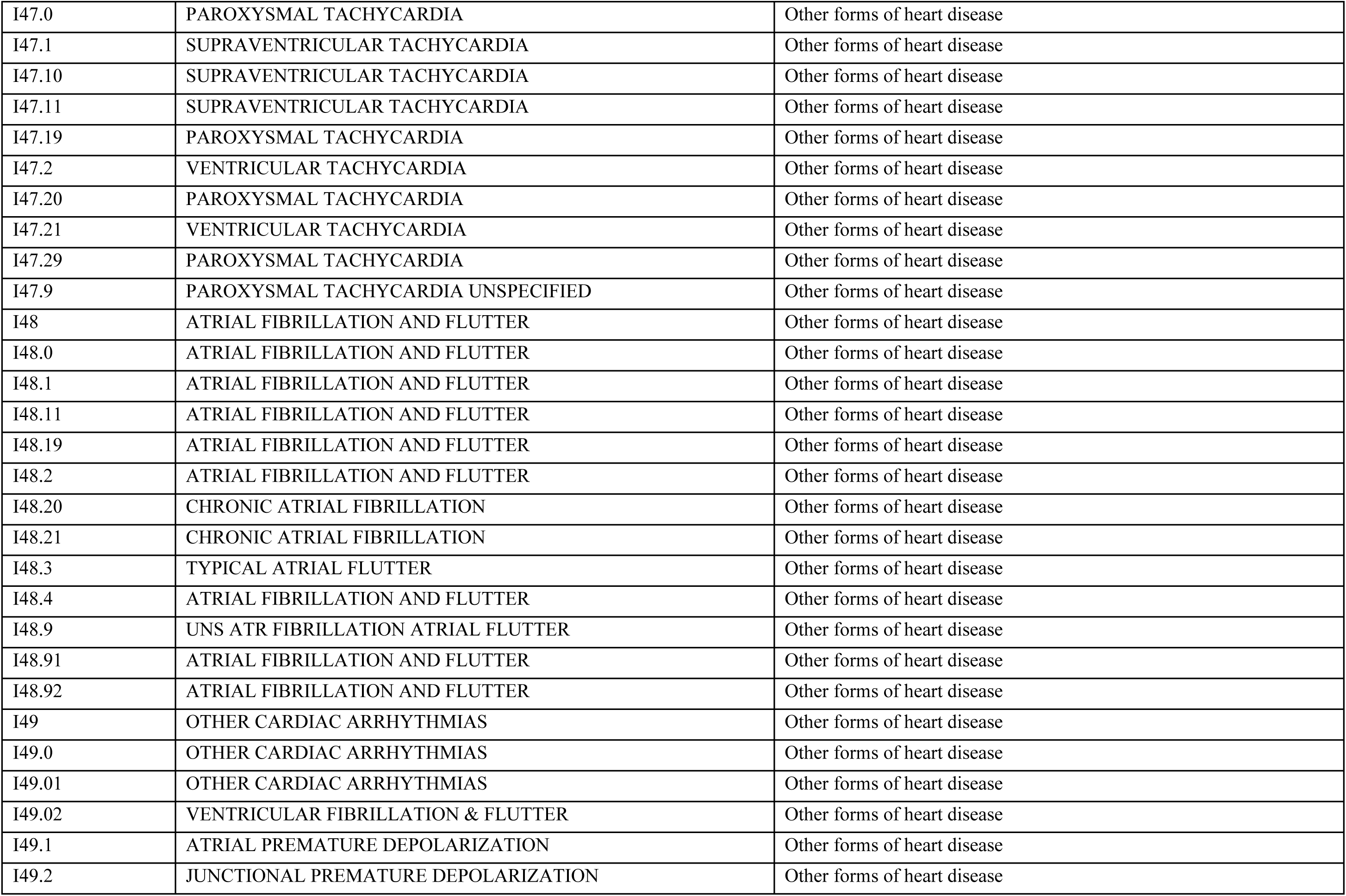
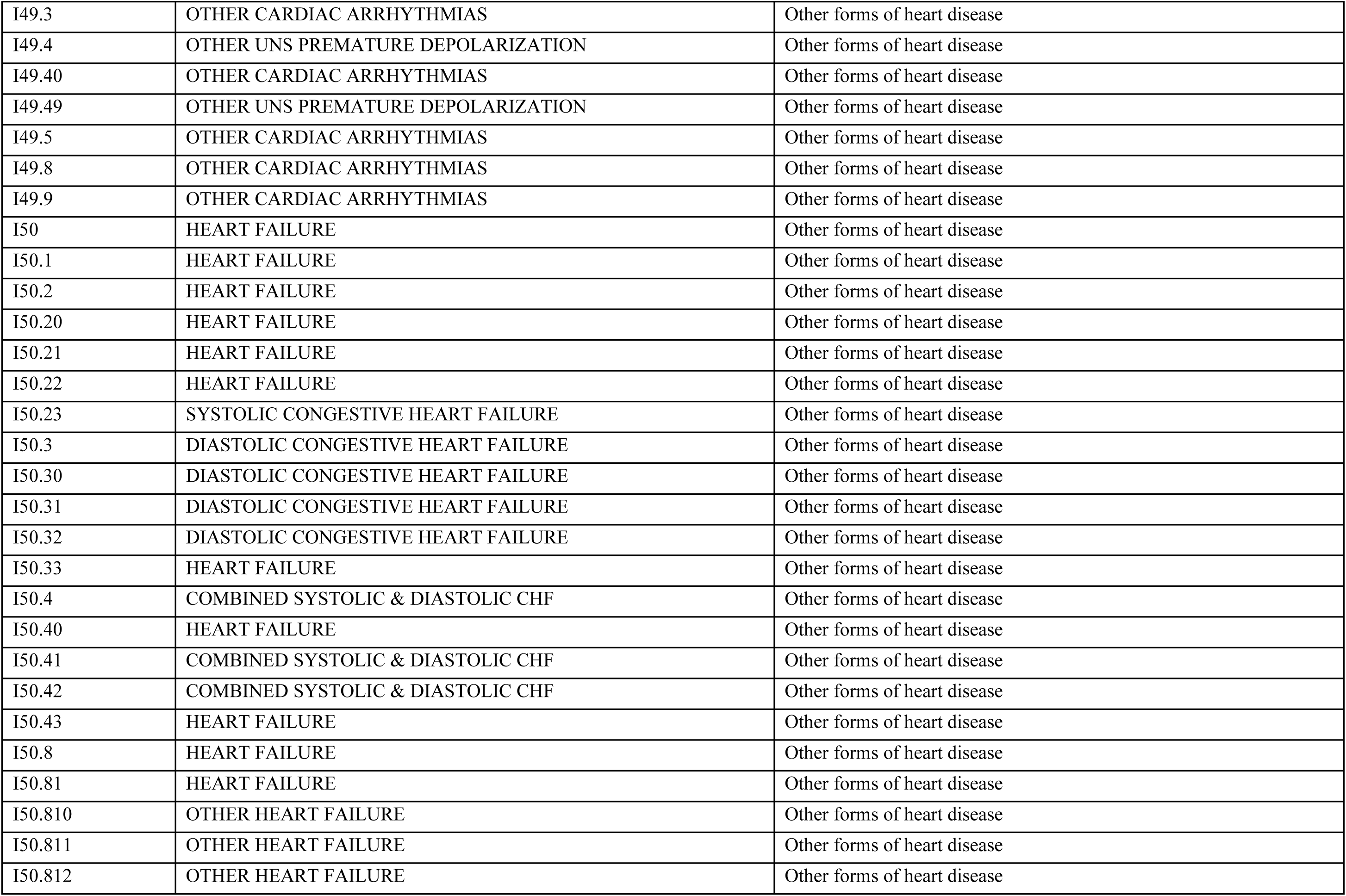
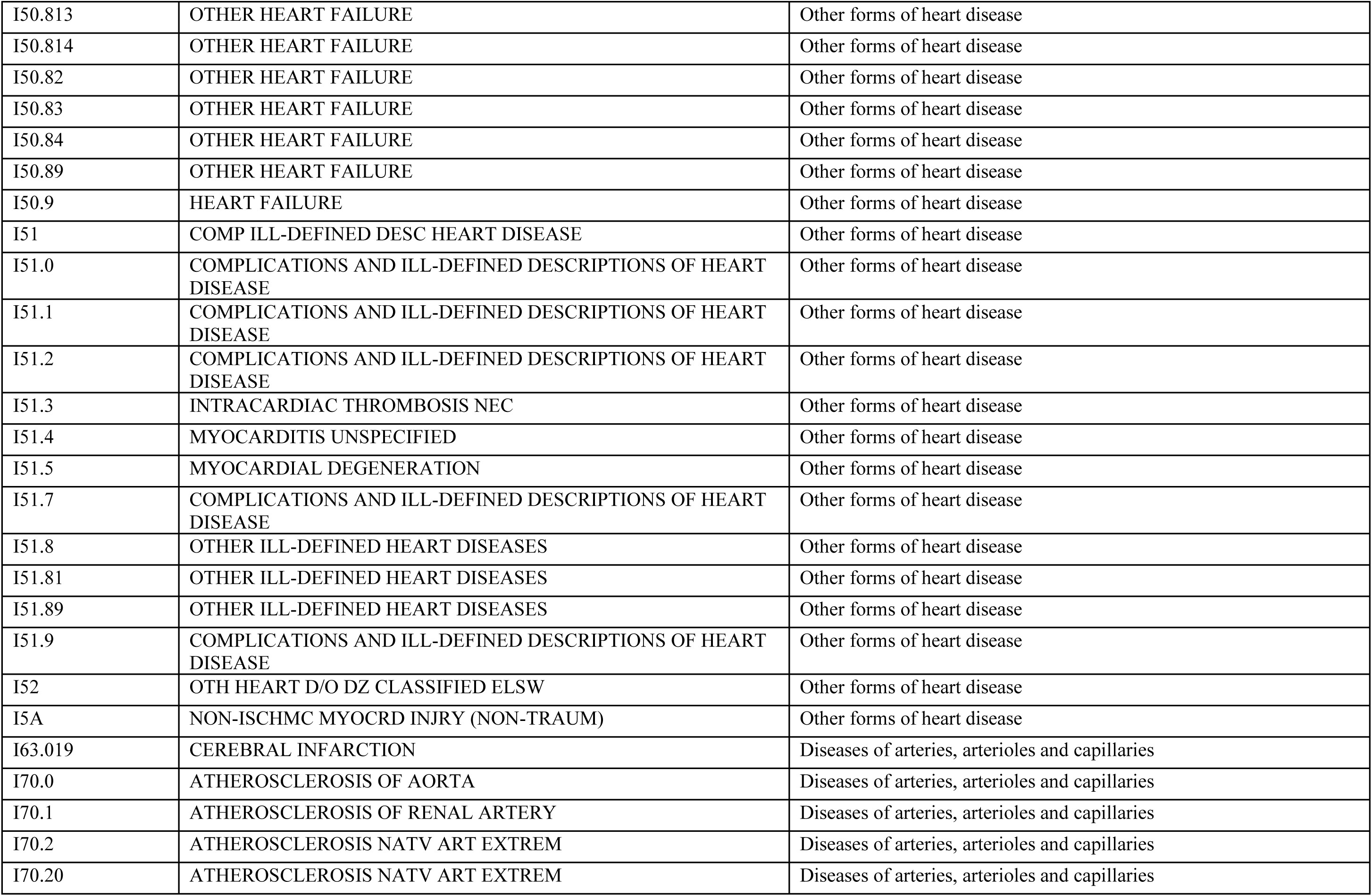
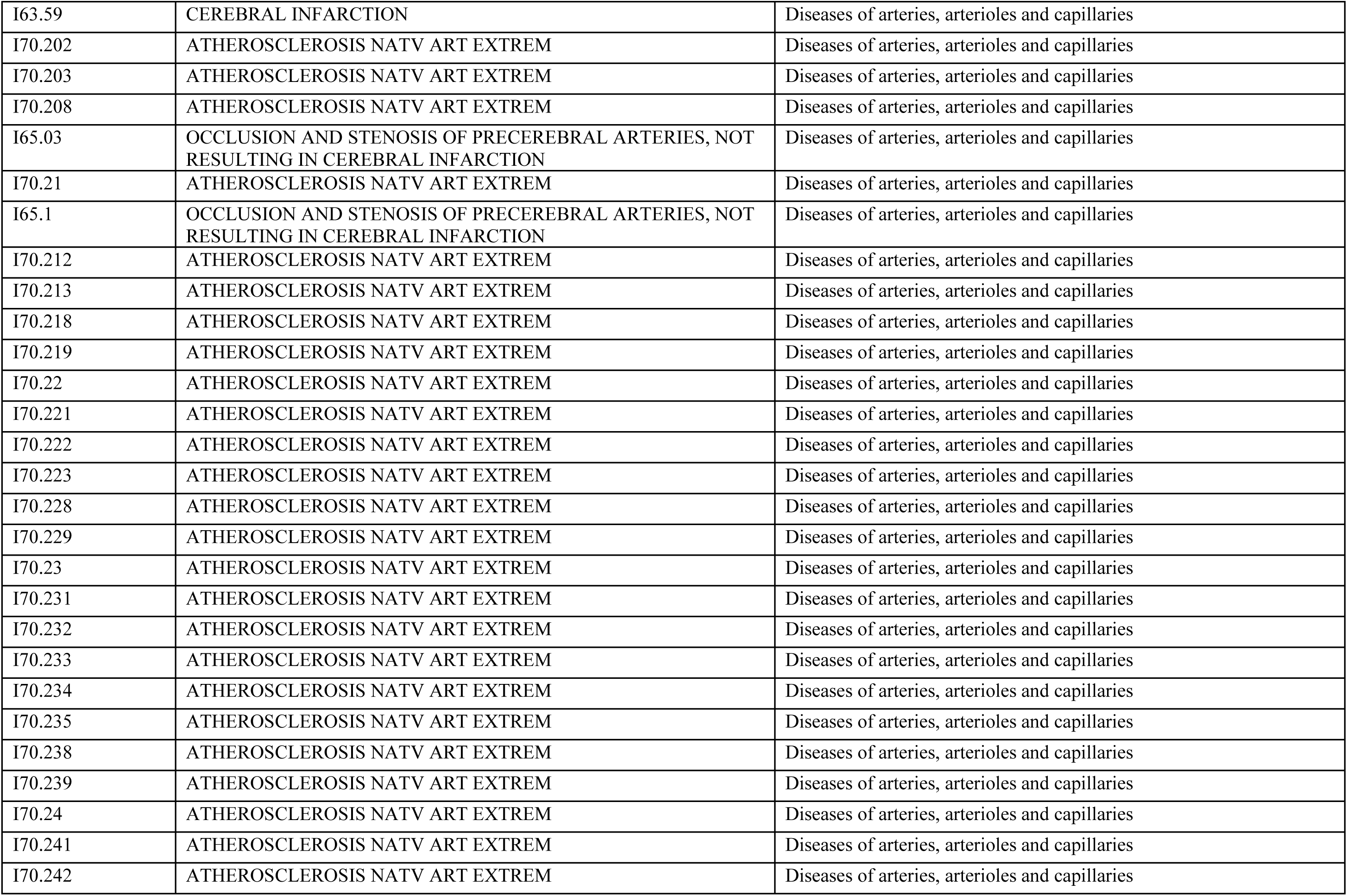
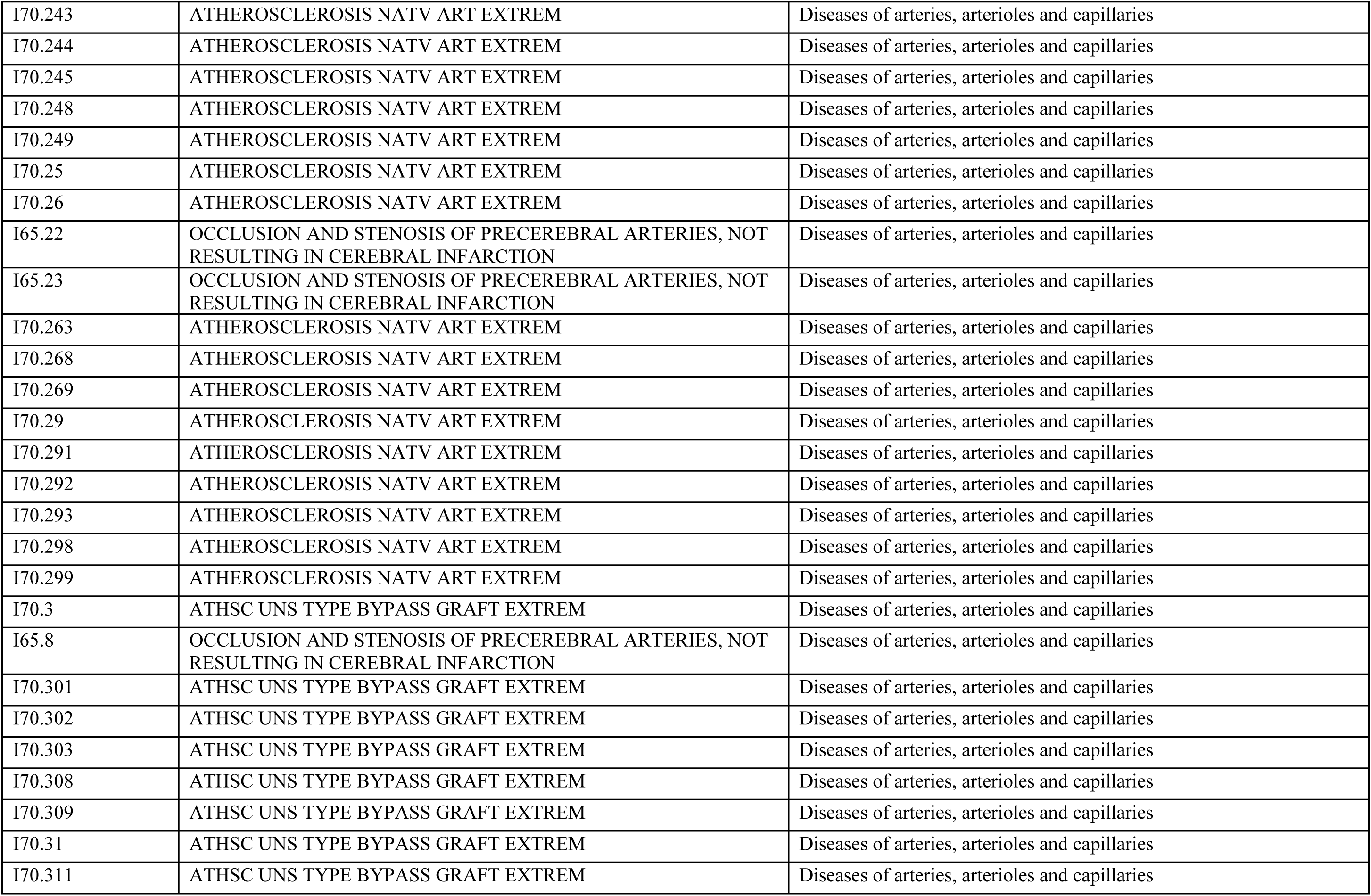
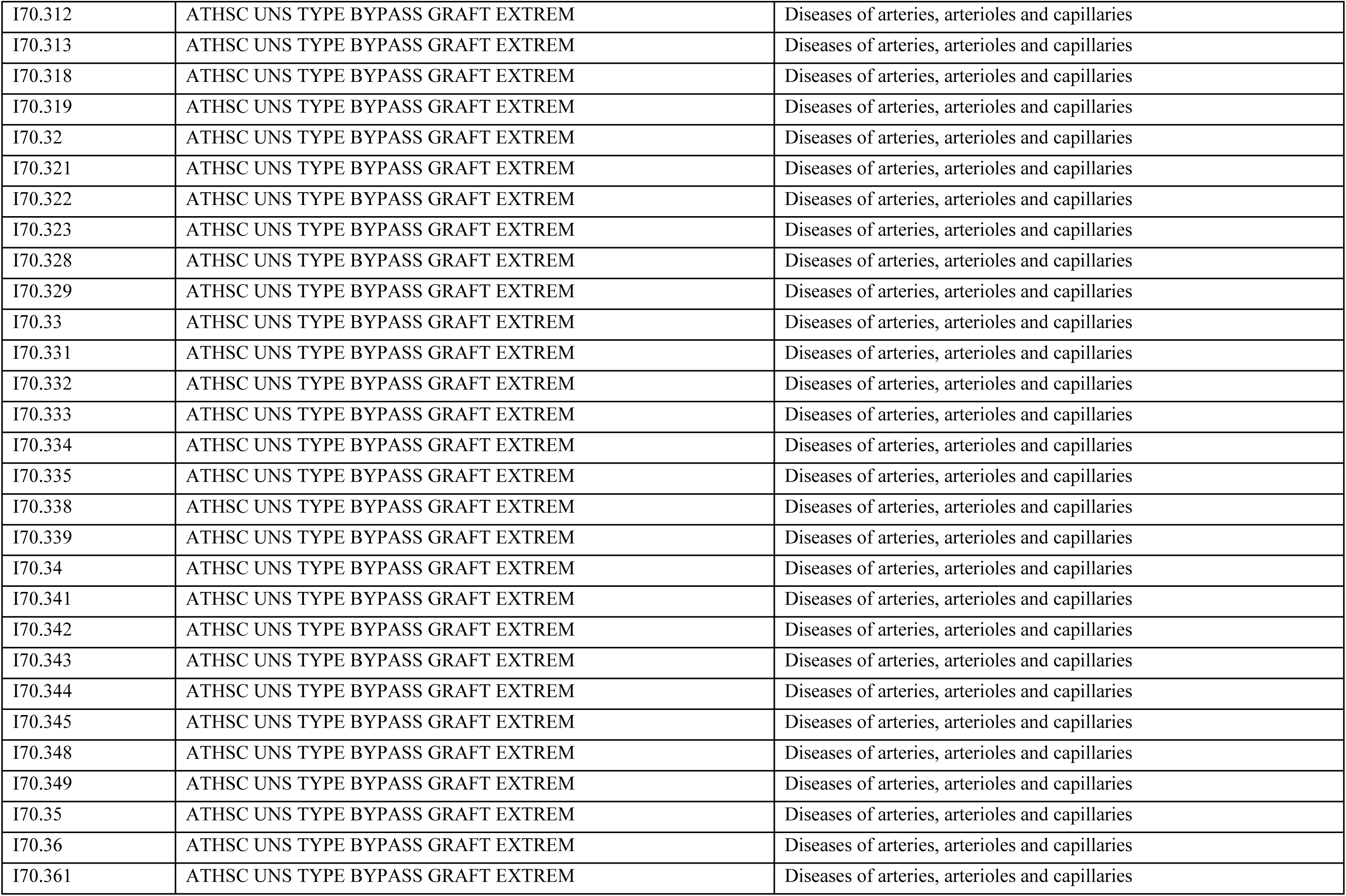
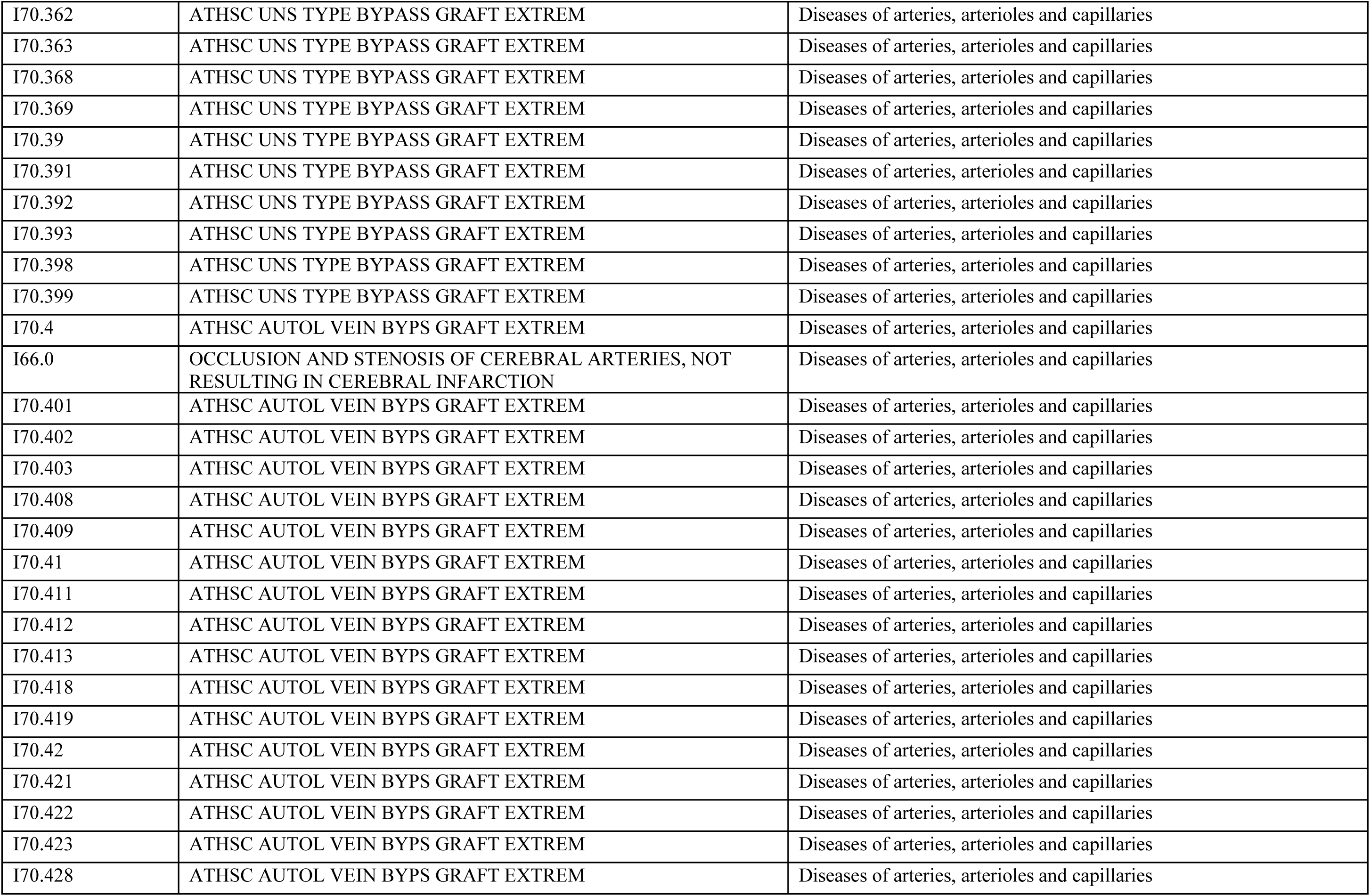
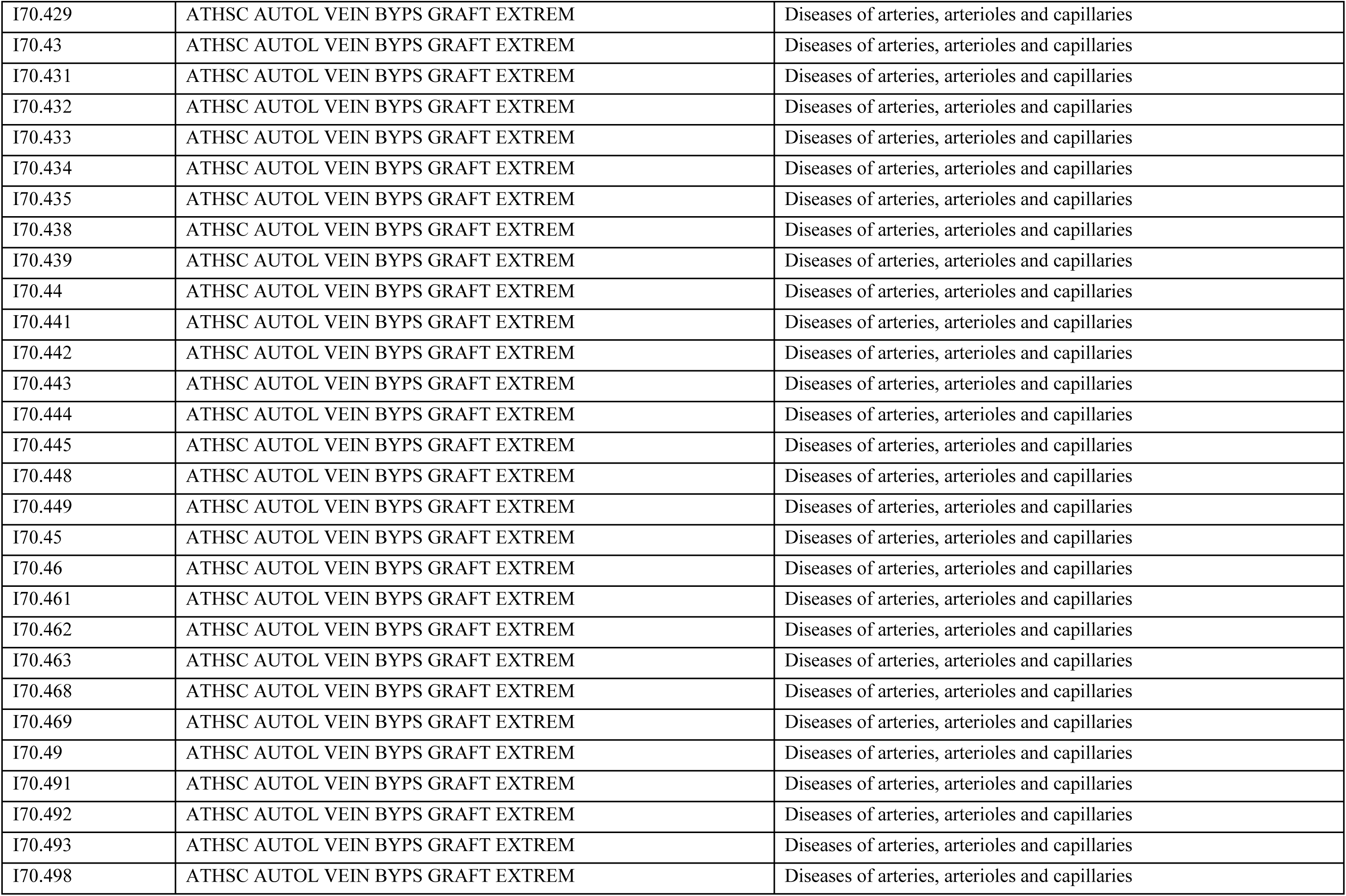
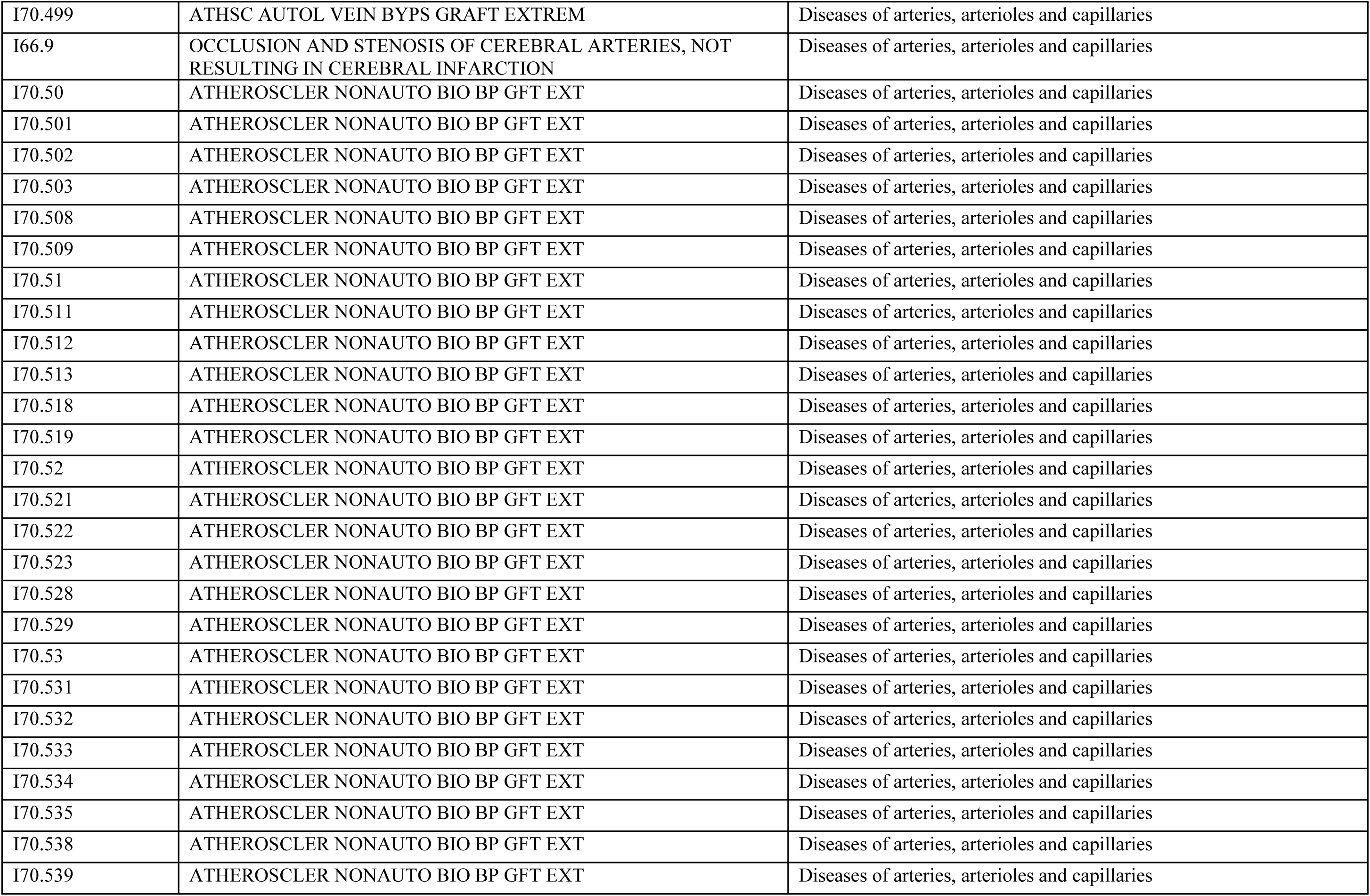
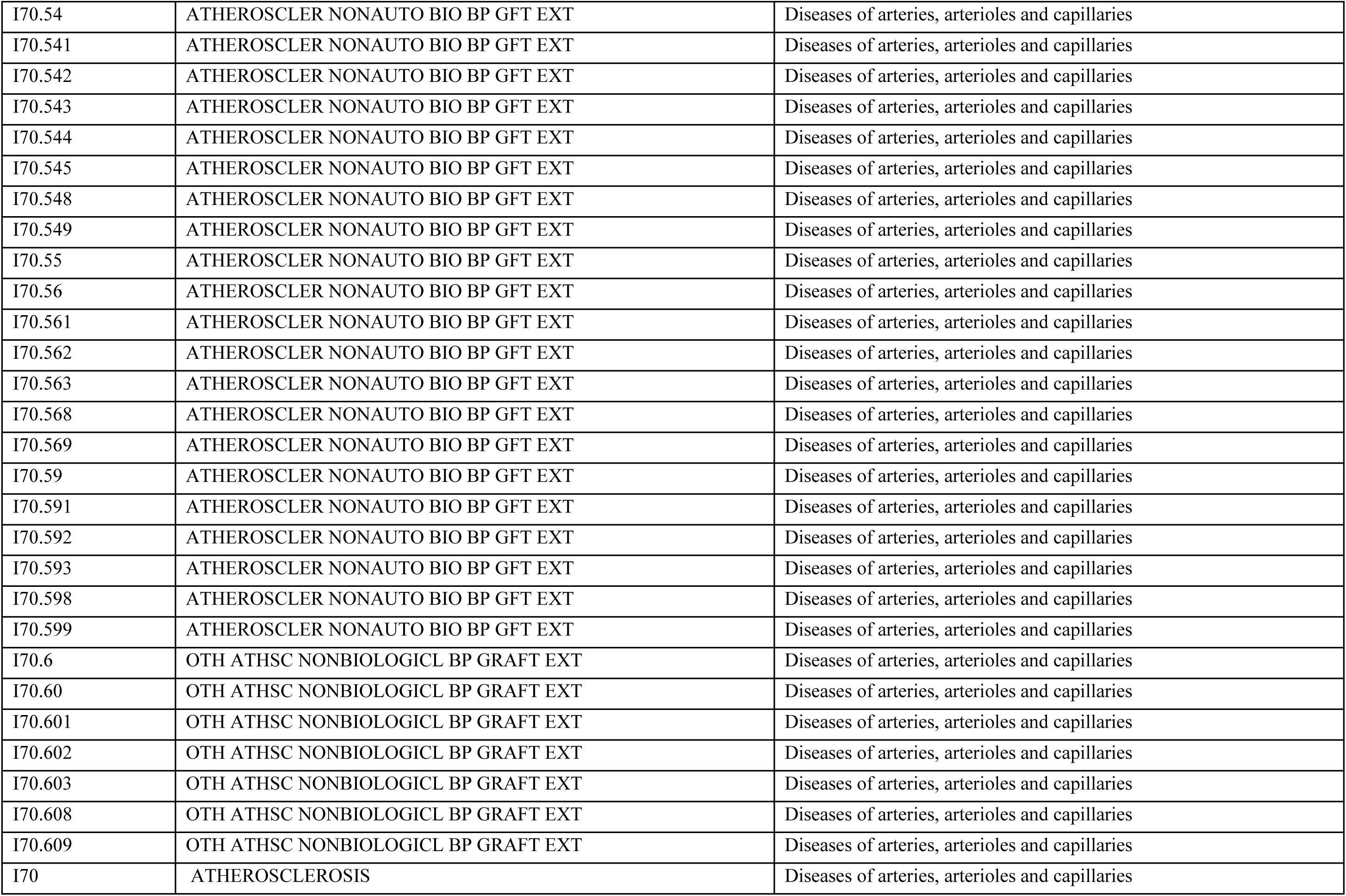
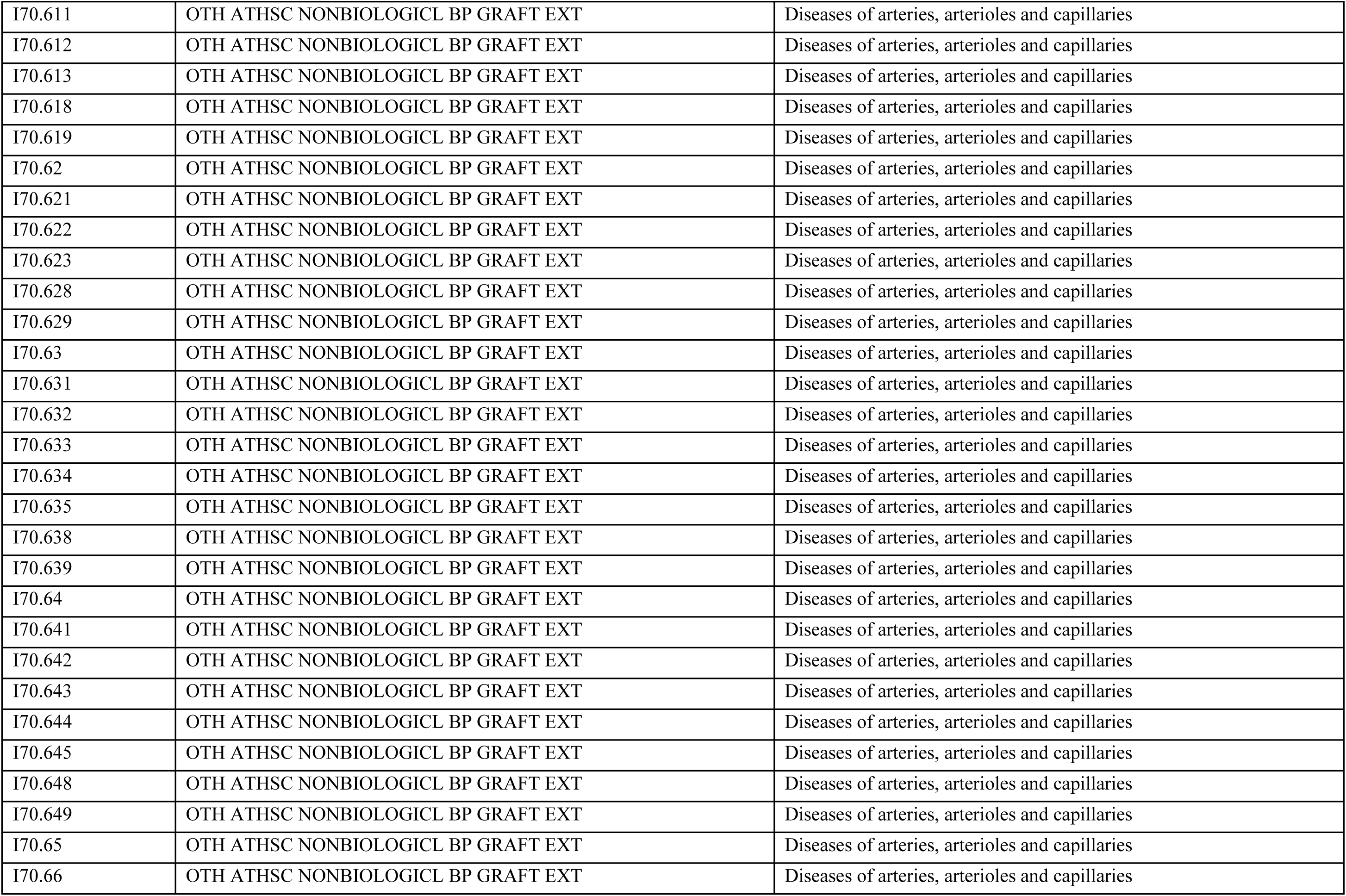
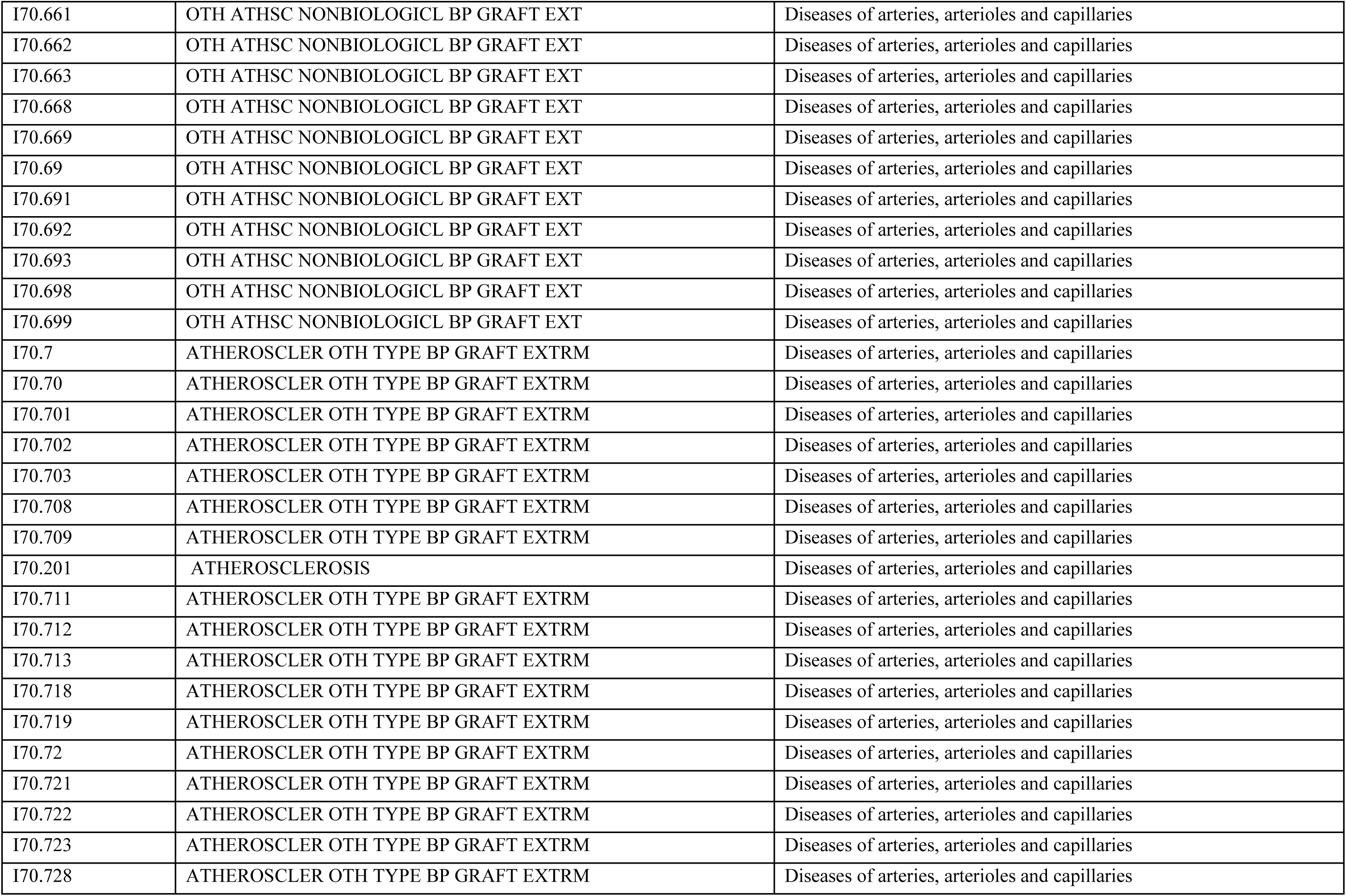
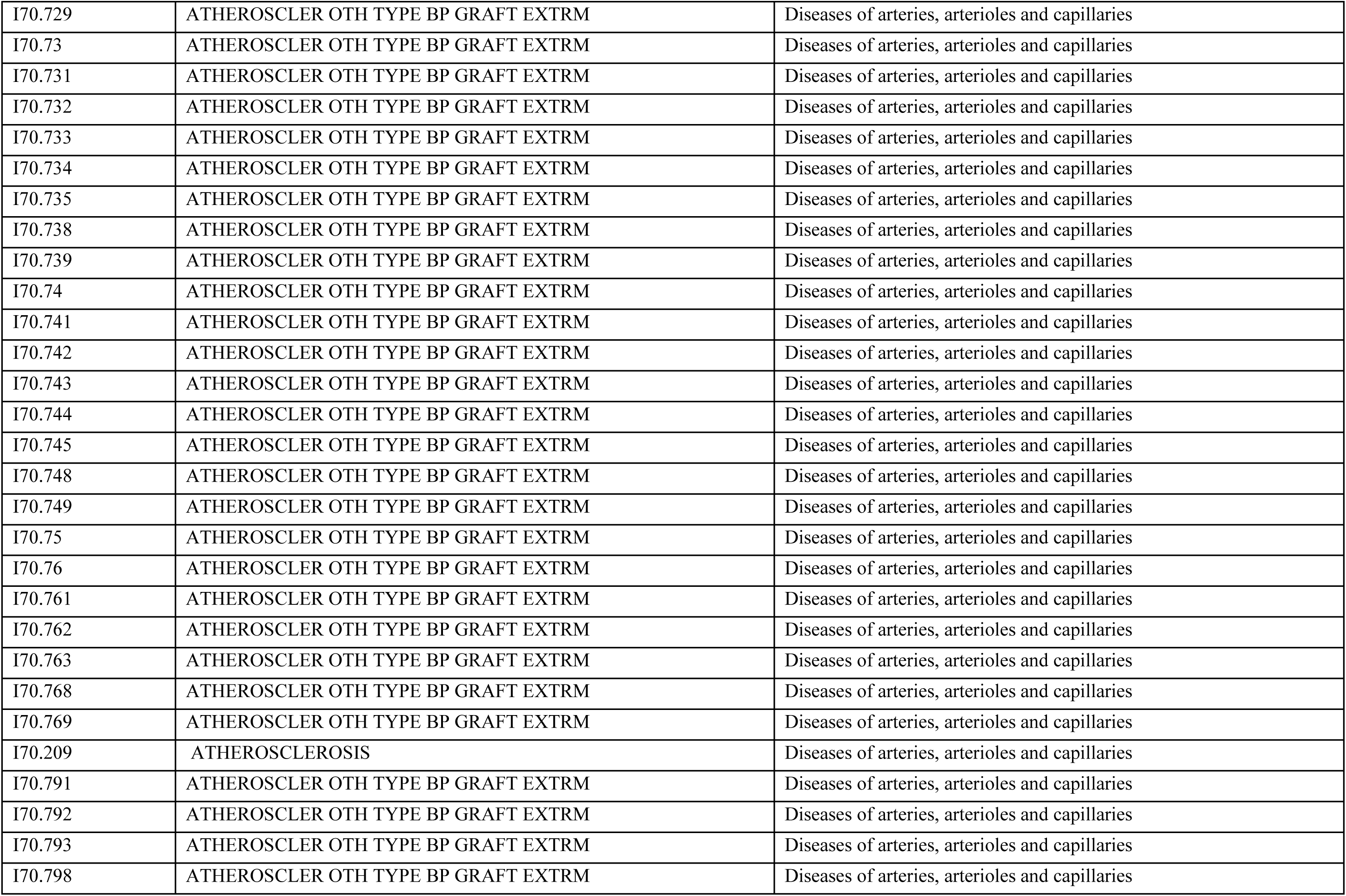
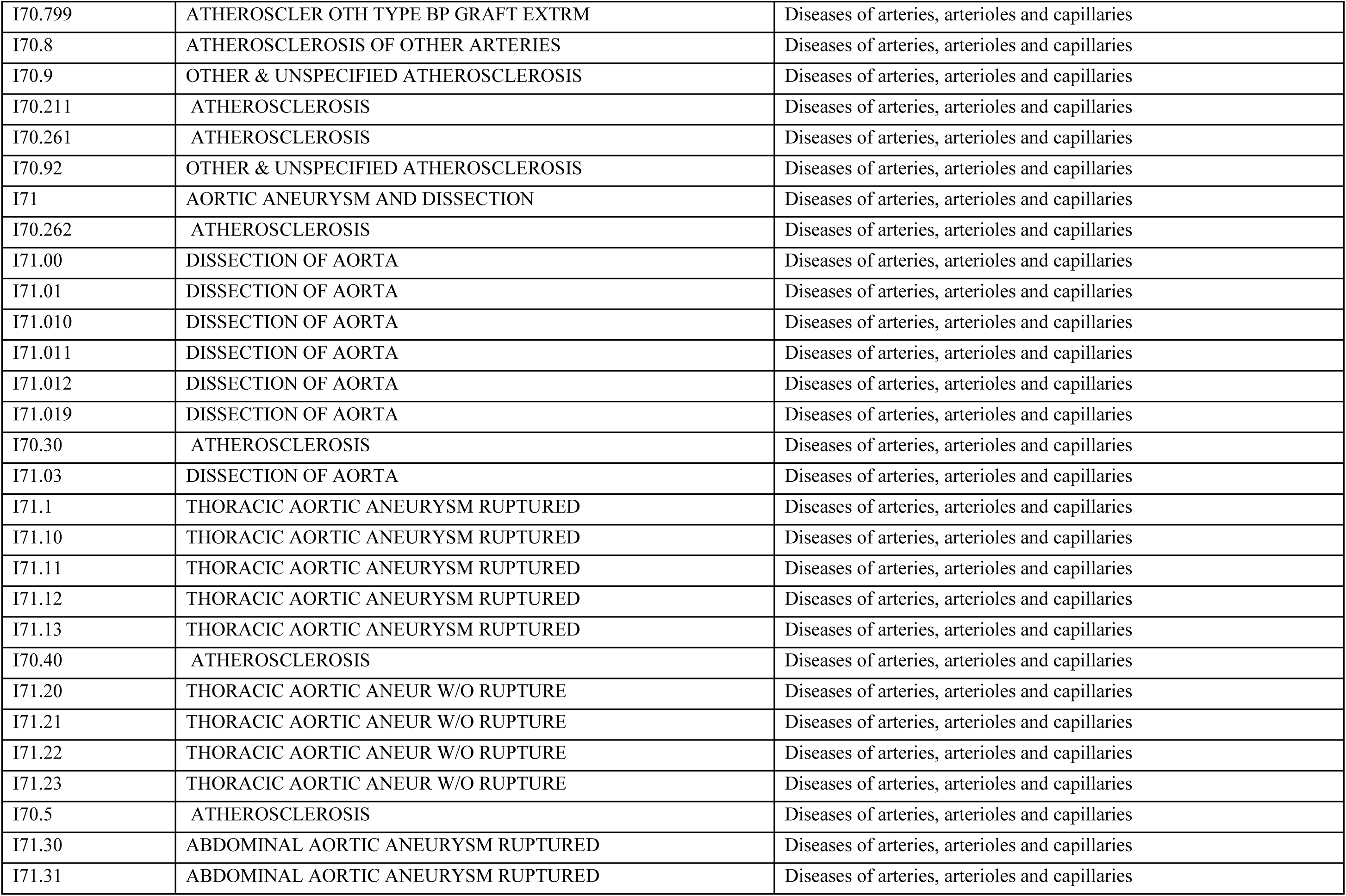
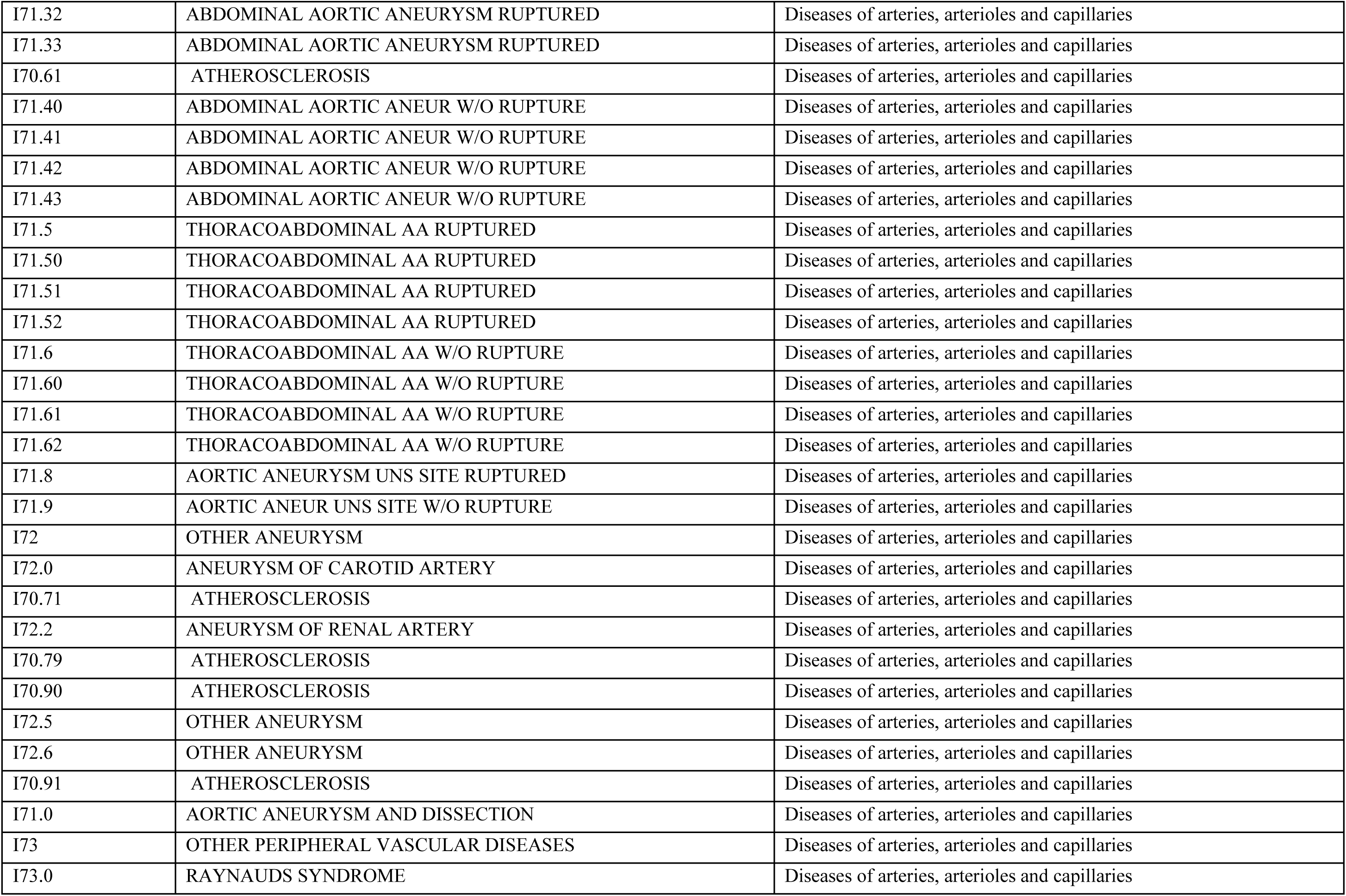
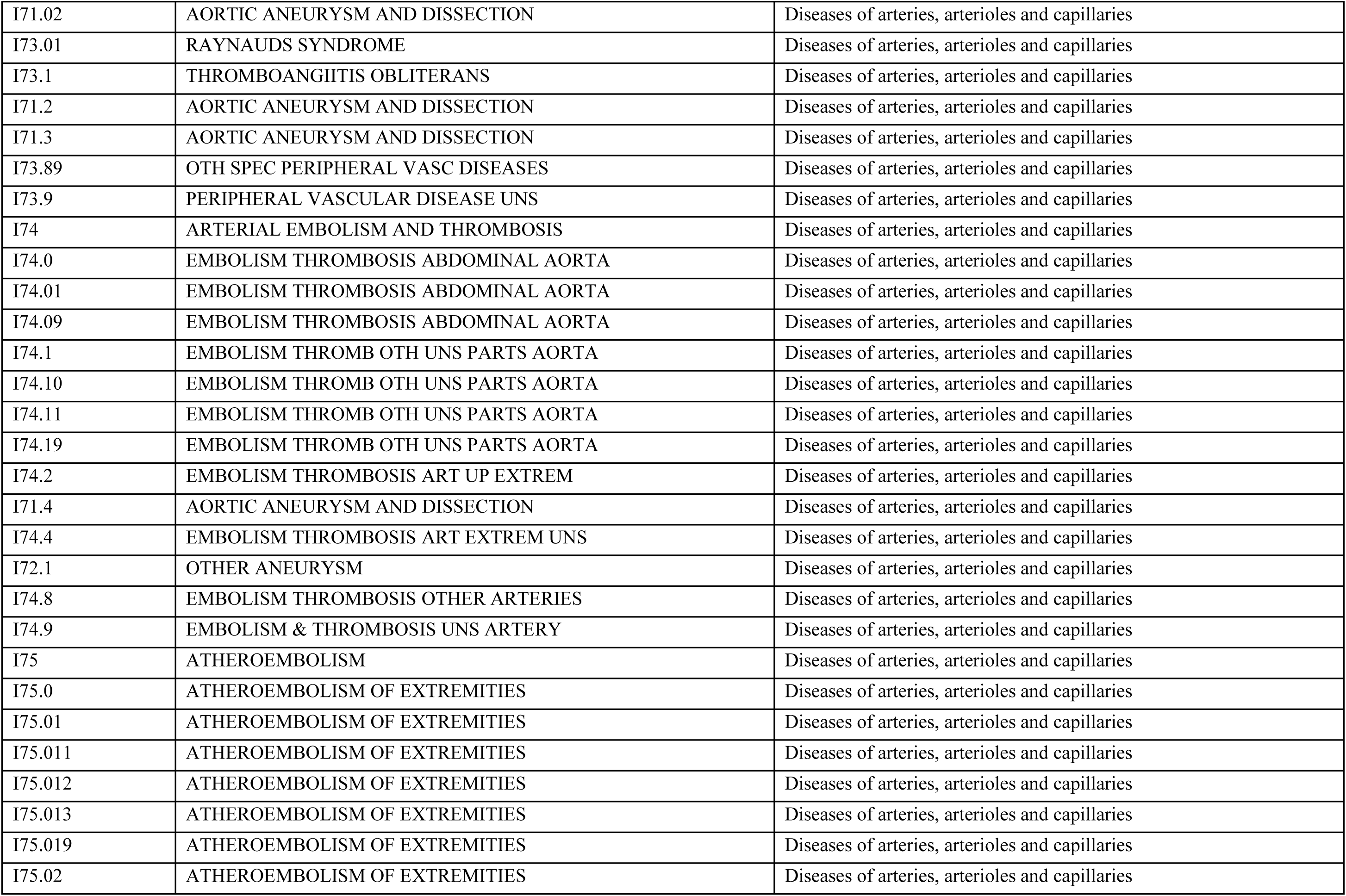
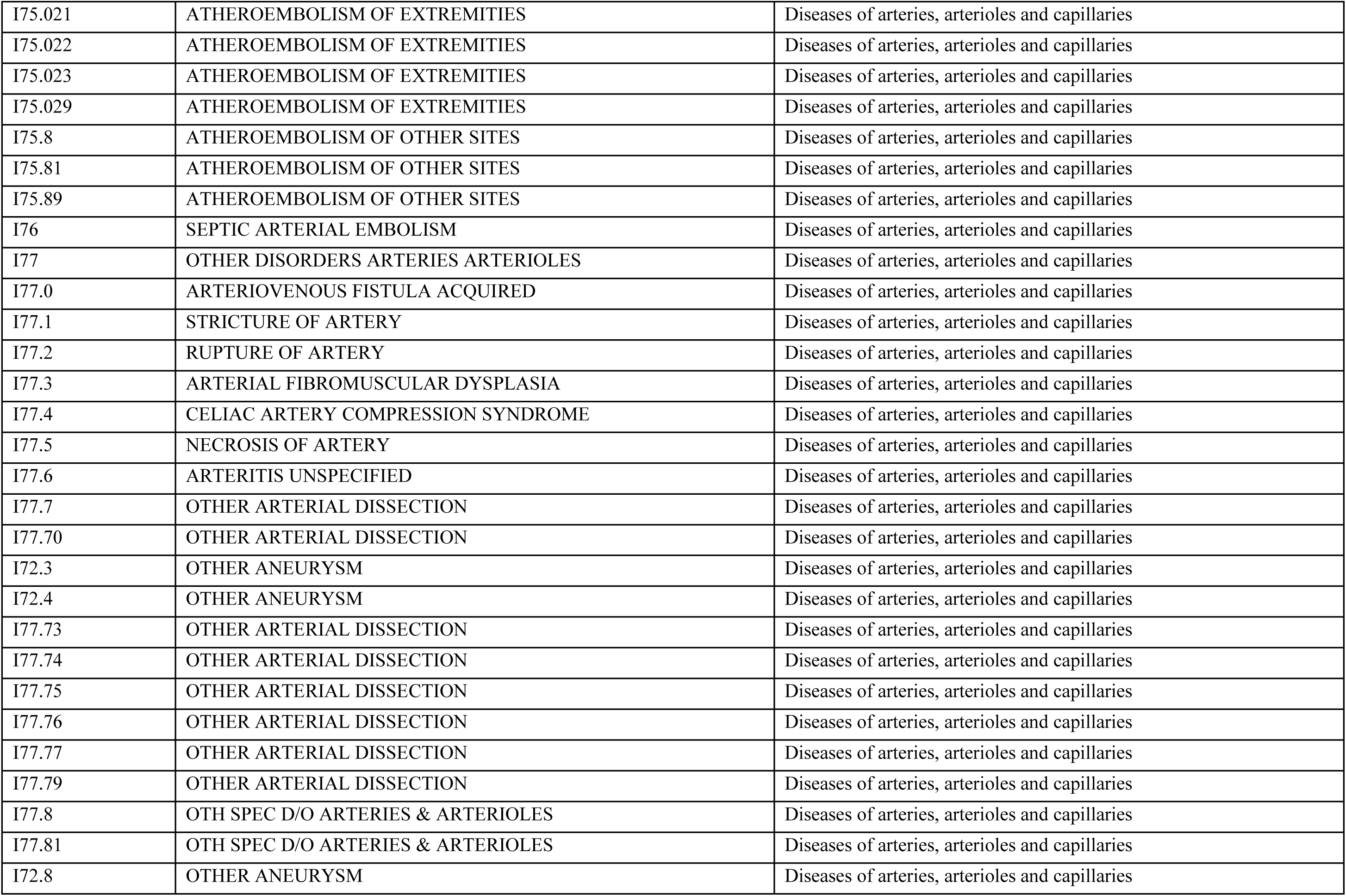
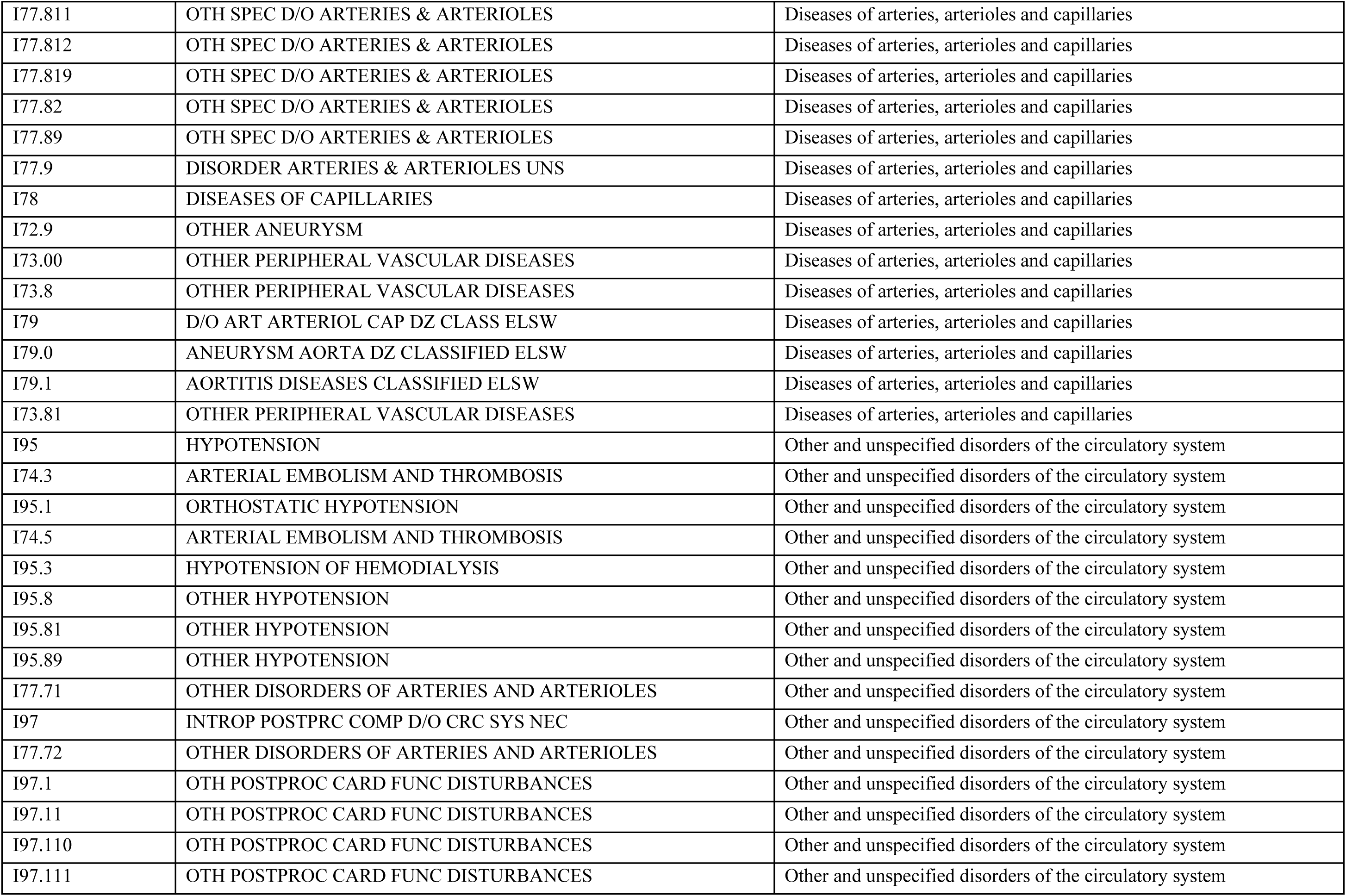
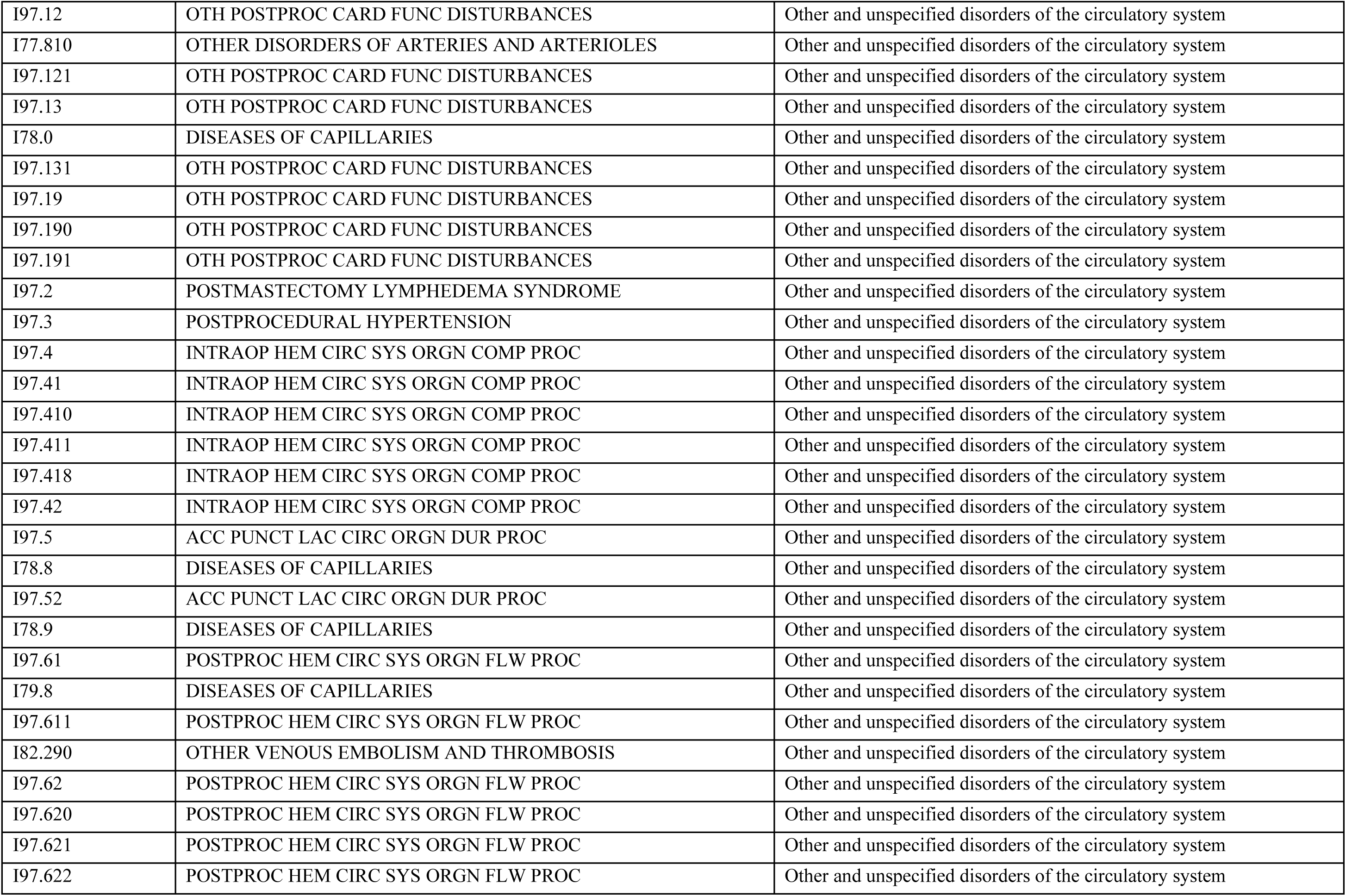
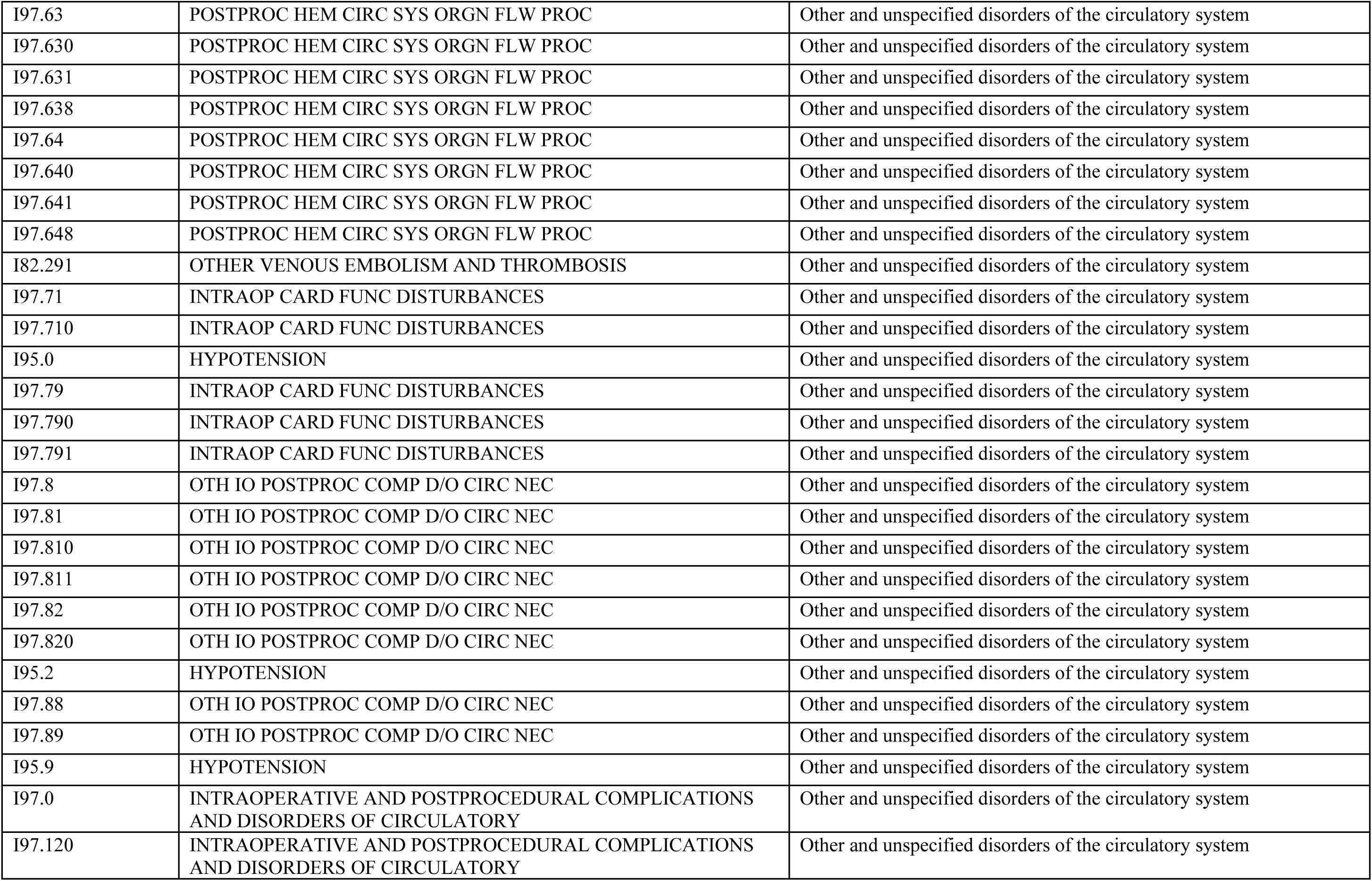
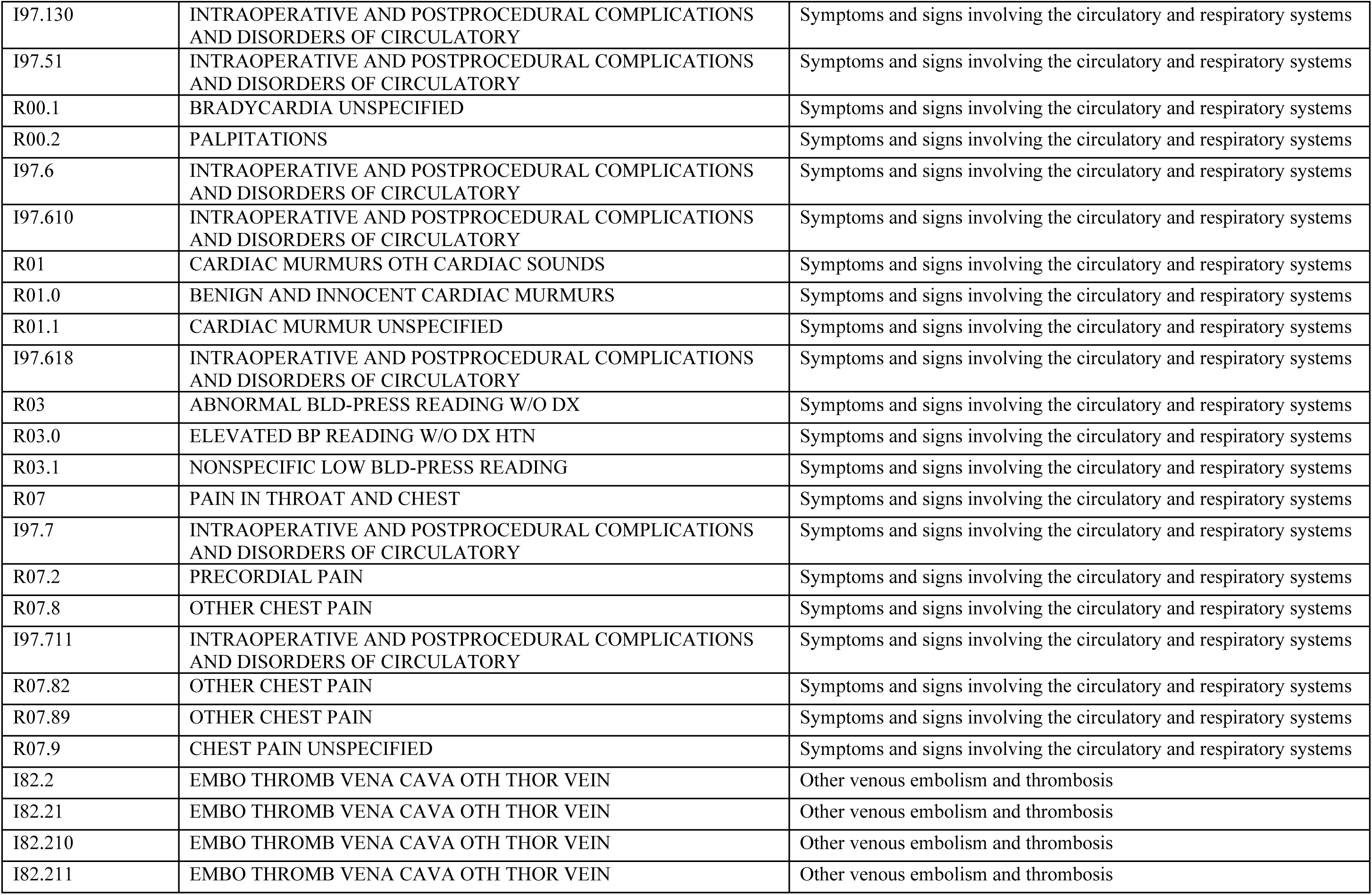
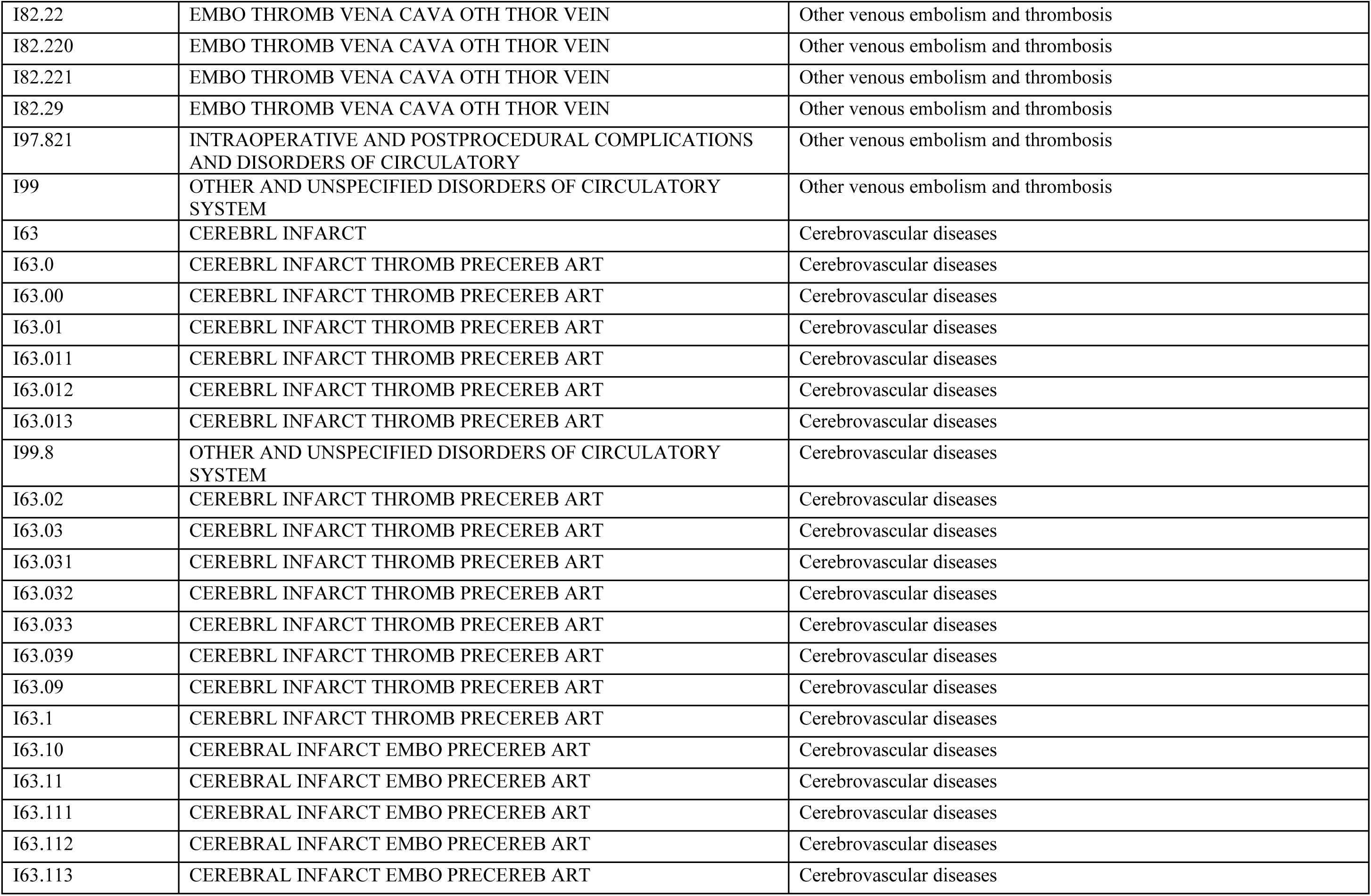
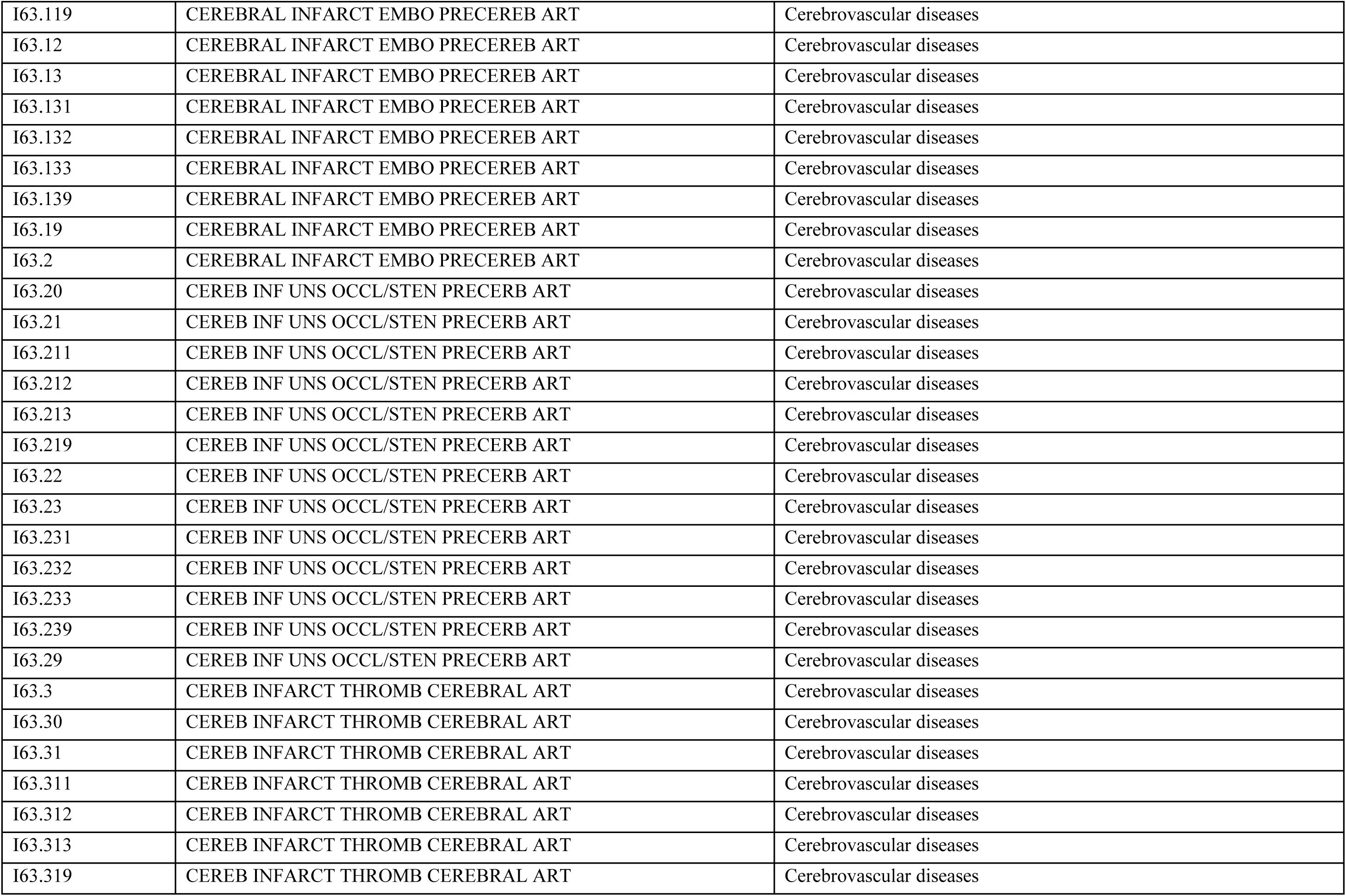
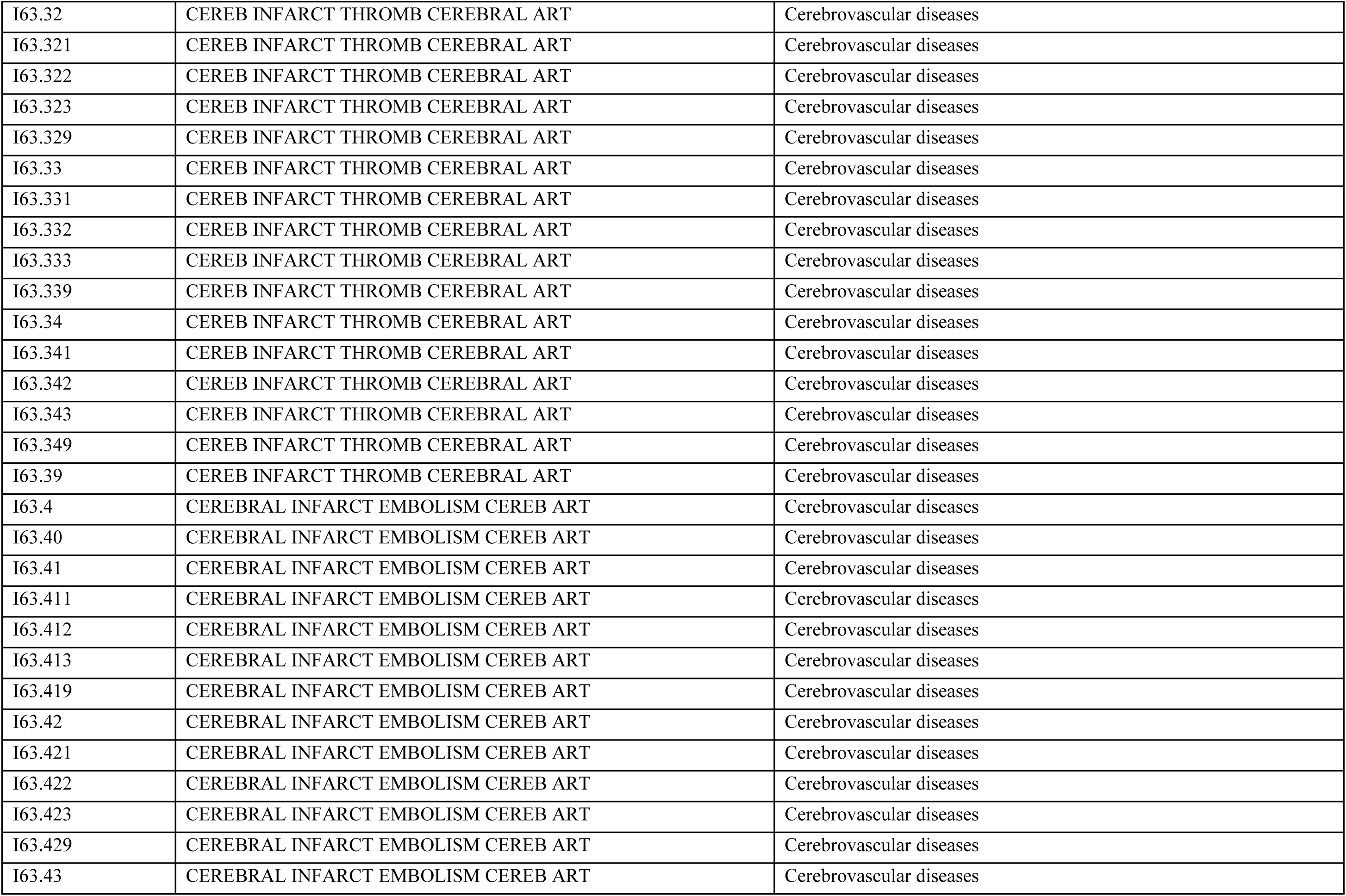
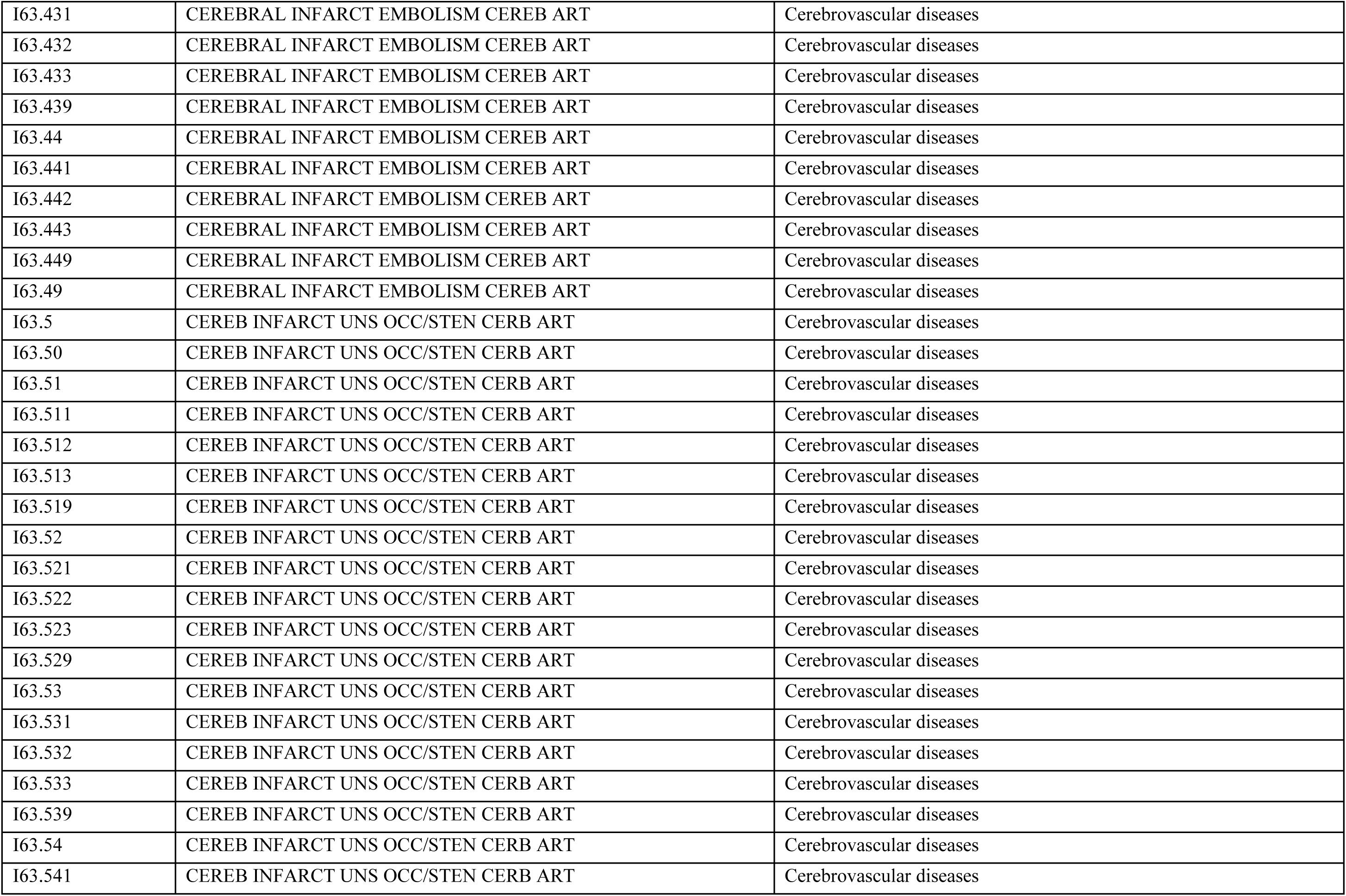
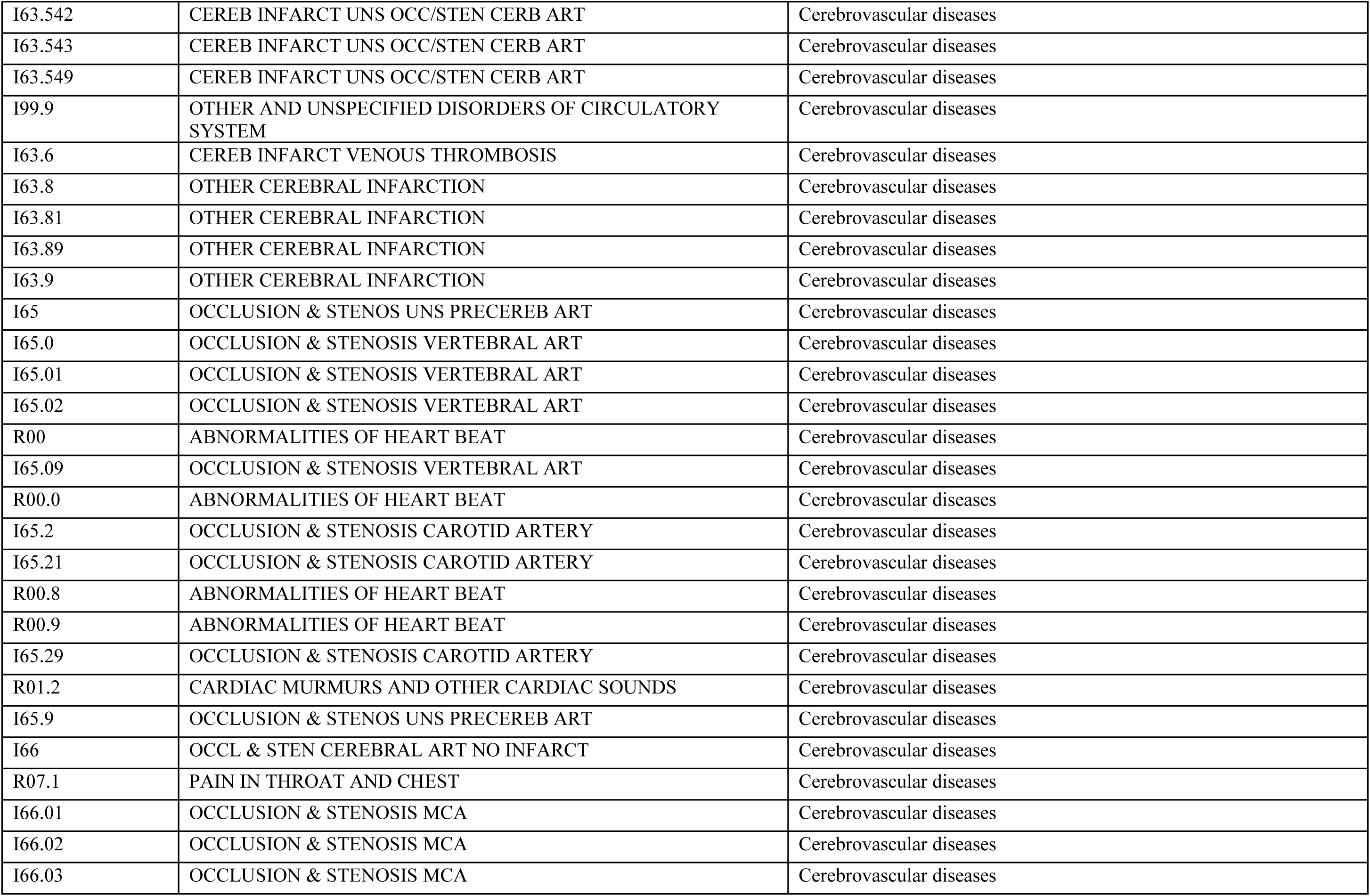
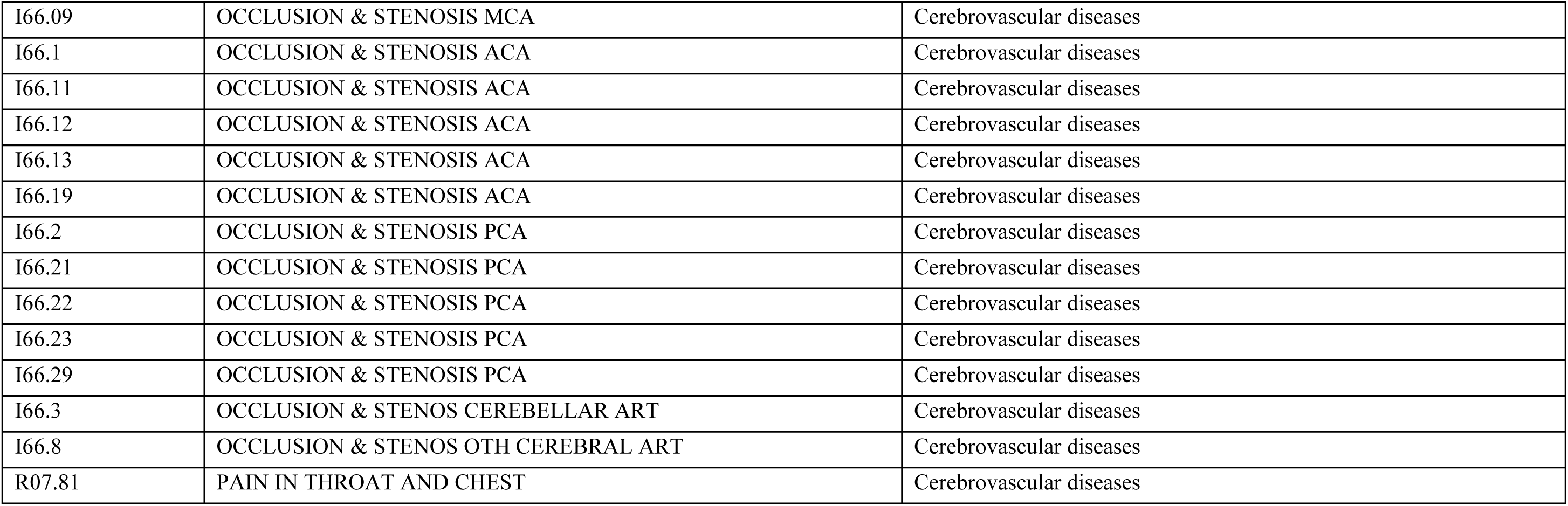
CV-related ICD-10-CM Codes Used in the Study.

**Supplemental Table 2.** Predictors of Time to Tafamidis Initiation by Socioeconomic Interactions.

| **Characteristic** | **HR (95% CI)** | ***P* value** |
| --- | --- | --- |
| **Sex** |  |  |
| Male | — | — |
| Female | 0.38 (0.32-0.44) | <0.001 |
| **Race** |  |  |
| White | — | — |
| Black | 1.05 (0.93-1.19) | 0.4 |
| **SDI** |  |  |
| ≤80 | — | — |
| >80 | 0.87 (0.77-0.99) | 0.041 |
| **ATTR-CM type** |  |  |
| ATTRwt-CM | — | — |
| ATTRv-CM | 1.12 (0.89-1.41) | 0.3 |
| **Year of diagnosis** |  |  |
| 2019-2020 | — | — |
| 2021-2022 | 1.22 (1.09-1.36) | <0.001 |
| 2023-2024 | 1.93 (1.72-2.17) | <0.001 |
| **Age at diagnosis** | 1.03 (1.03-1.04) | <0.001 |
| **US region** |  |  |
| Northeast | — | — |
| Midwest | 0.95 (0.86-1.05) | 0.3 |
| South | 0.89 (0.79-0.99) | 0.036 |
| West | 0.75 (0.65-0.88) | <0.001 |
| **Insurance type** |  |  |
| Medicare | — | — |
| Other/Unknown | 1.29 (1.12-1.50) | <0.001 |
| **Disease severity** |  |  |
| None | — | — |
| Mild or minor | 1.61 (1.46-1.78) | <0.001 |
| Moderate or severe | 1.50 (1.31-1.72) | <0.001 |
| **Comorbidity** |  |  |
| CKD | 0.82 (0.75-0.90) | <0.001 |
| COPD | 0.77 (0.69-0.86) | <0.001 |
| Coronary artery disease | 1.05 (0.96-1.15) | 0.2 |
| Diabetes mellitus | 0.76 (0.70-0.84) | <0.001 |
| Diastolic heart failure | 1.38 (1.26-1.51) | <0.001 |
| Hypertension | 0.81 (0.72-0.92) | <0.001 |
| Obesity | 0.87 (0.79-0.97) | 0.011 |
| Stroke | 0.49 (0.43-0.55) | **<0.001** |
| Systolic heart failure | 1.17 (1.07-1.28) | **<0.001** |
| **Sex\*race** |  |  |
| Female*Black | 1.91 (1.54-2.38) | **<0.001** |
| **Sex\*SDI** |  |  |
| Female*SDI >80 | 1.13 (0.89-1.43) | 0.3 |
| **Race\*ATTR-CM type** |  |  |
| Black*ATTRv-CM | 1.65 (1.24-2.20) | **<0.001** |

ATTR-CM = transthyretin amyloid cardiomyopathy; ATTRv-CM = variant transthyretin amyloid cardiomyopathy; ATTRwt-CM = wild-type transthyretin amyloid cardiomyopathy; CKD = chronic kidney disease; COPD = chronic obstructive pulmonary disease; HR = hazard ratio; SDI = Social Deprivation Index.

**Supplemental Table 3.** Pairwise Contrasts of Sex and Race for Time to Tafamidis Initiation.

| **Contrast** | **HR (95% CI)** | **P value** |
| --- | --- | --- |
| **White female vs White male (reference)** |  |  |
| Overall | 0.39 (0.33-0.45) | **<0.001** |
| ATTRwt-CM | 0.39 (0.33-0.45) | **<0.001** |
| ATTRv-CM | 0.39 (0.33-0.45) | **<0.001** |
| SDI ≤80 | 0.38 (0.32-0.44) | **<0.001** |
| SDI >80 | 0.43 (0.33-0.55) | **<0.001** |
| **Black male vs White male (reference)** |  |  |
| Overall | 1.10 (0.98-1.23) | 0.12 |
| ATTRwt-CM | 1.05 (0.93-1.19) | 0.42 |
| ATTRv-CM | 1.74 (1.32-2.30) | **<0.001** |
| SDI ≤80 | 1.10 (0.98-1.23) | 0.12 |
| SDI >80 | 1.10 (0.98-1.23) | 0.12 |
| **Black male vs White female (reference)** |  |  |
| Overall | 2.83 (2.38-3.37) | **<0.001** |
| ATTRwt-CM | 2.71 (2.27-3.24) | **<0.001** |
| ATTRv-CM | 4.49 (3.30-6.10) | **<0.001** |
| SDI ≤80 | 2.91 (2.43-3.48) | **<0.001** |
| SDI >80 | 2.58 (1.99-3.34) | **<0.001** |
| **Black female vs White male (reference)** |  |  |
| Overall | 0.81 (0.71-0.93) | 0.003 |
| ATTRwt-CM | 0.78 (0.68-0.90) | **<0.001** |
| ATTRv-CM | 1.29 (0.97-1.72) | 0.08 |
| SDI ≤80 | 0.79 (0.68-0.93) | 0.003 |
| SDI >80 | 0.89 (0.73-1.10) | 0.29 |
| **Black female vs White female (reference)** |  |  |
| Overall | 2.10 (1.73-2.54) | **<0.001** |
| ATTRwt-CM | 2.01 (1.66-2.45) | **<0.001** |
| ATTRv-CM | 3.33 (2.43-4.57) | **<0.001** |
| SDI ≤80 | 2.10 (1.73-2.54) | **<0.001** |
| SDI >80 | 2.10 (1.73-2.54) | **<0.001** |
| **Black female vs Black male (reference)** |  |  |
| Overall | 0.74 (0.64-0.86) | **<0.001** |
| ATTRwt-CM | 0.74 (0.64-0.86) | **<0.001** |
| ATTRv-CM | 0.74 (0.64-0.86) | **<0.001** |
| SDI ≤80 | 0.72 (0.61-0.86) | **<0.001** |
| SDI >80 | 0.81 (0.67-1.00) | 0.05 |

ATTRv-CM = variant transthyretin amyloid cardiomyopathy; ATTRwt-CM = wild-type transthyretin amyloid cardiomyopathy; HR = hazard ratio; SDI = Social Deprivation Index.

**Supplemental Table 4.**
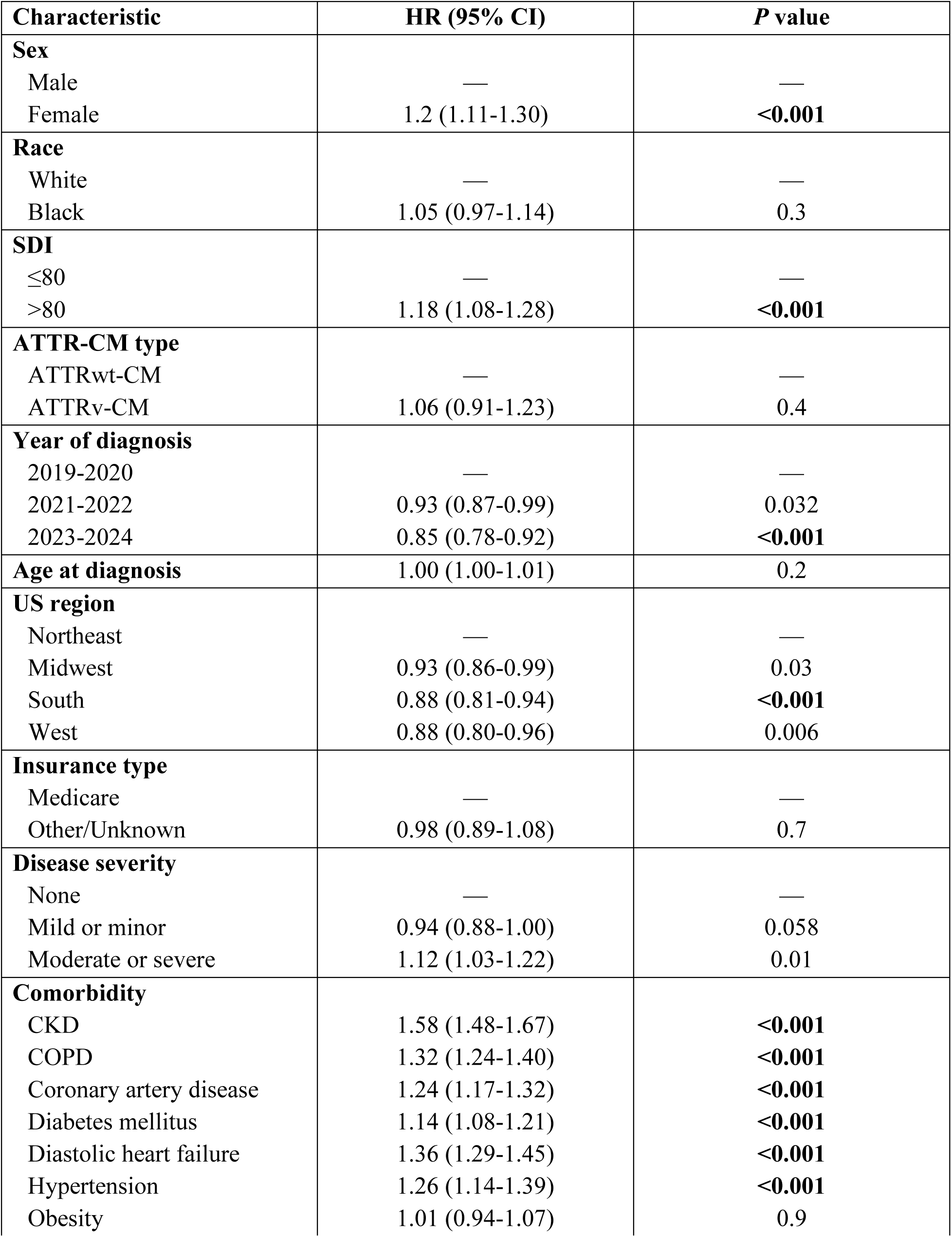
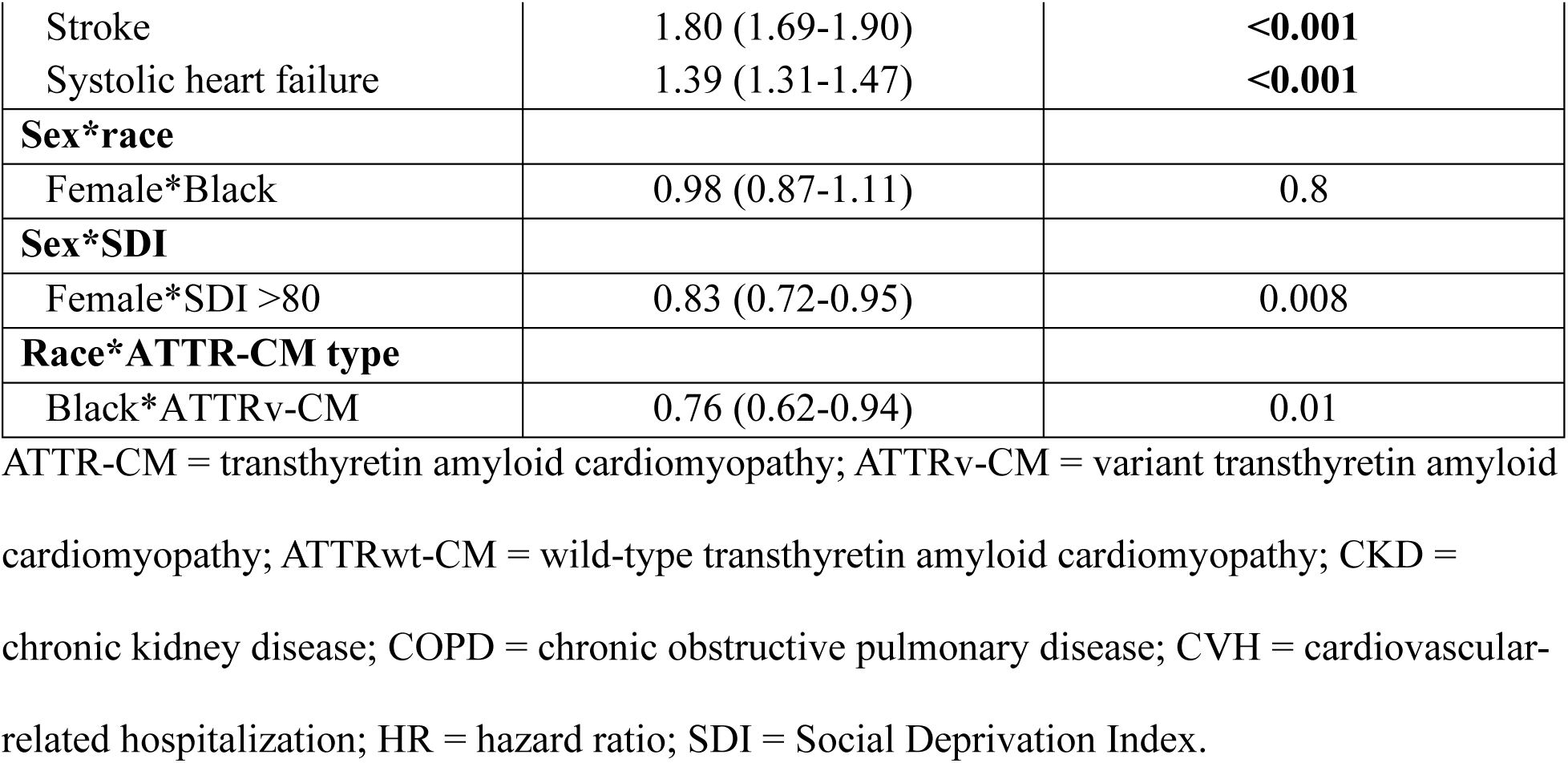
Predictors of Time to CVH or Death by Socioeconomic Interactions.

**Supplemental Table 5.** Pairwise Contrasts of Sex and Race for Time to CVH or Death by SDI Score.

| **Contrast** | **HR (95% CI)** | **P value** |
| --- | --- | --- |
| **White female vs White male (reference)** |  |  |
| Overall | 1.15 (1.07-1.25) | **<0.001** |
| ATTRwt-CM | 1.15 (1.07-1.25) | **<0.001** |
| ATTRv-CM | 1.15 (1.07-1.25) | **<0.001** |
| SDI ≤80 | 1.20 (1.11-1.30) | **<0.001** |
| SDI >80 | 1.00 (0.87-1.15) | 0.98 |
| **Black male vs White male (reference)** |  |  |
| Overall | 1.03 (0.95-1.11) | 0.54 |
| ATTRwt-CM | 1.05 (0.97-1.14) | 0.26 |
| ATTRv-CM | 0.80 (0.65-0.98) | 0.03 |
| SDI ≤80 | 1.03 (0.95-1.11) | 0.54 |
| SDI >80 | 1.03 (0.95-1.11) | 0.54 |
| **Black male vs White female (reference)** |  |  |
| Overall | 0.89 (0.81-0.98) | 0.01 |
| ATTRwt-CM | 0.91 (0.83-1.00) | 0.05 |
| ATTRv-CM | 0.69 (0.56-0.86) | **<0.001** |
| SDI ≤80 | 0.85 (0.77-0.94) | 0.002 |
| SDI >80 | 1.03 (0.89-1.18) | 0.70 |
| **Black female vs White male (reference)** |  |  |
| Overall | 1.16 (1.07-1.26) | **<0.001** |
| ATTRwt-CM | 1.19 (1.09-1.29) | **<0.001** |
| ATTRv-CM | 0.91 (0.74-1.11) | 0.35 |
| SDI ≤80 | 1.21 (1.10-1.33) | **<0.001** |
| SDI >80 | 1.00 (0.89-1.14) | 0.94 |
| **Black female vs White female (reference)** |  |  |
| Overall | 1.01 (0.91-1.11) | 0.90 |
| ATTRwt-CM | 1.03 (0.93-1.14) | 0.57 |
| ATTRv-CM | 0.79 (0.64-0.97) | 0.03 |
| SDI ≤80 | 1.01 (0.91-1.11) | 0.90 |
| SDI >80 | 1.01 (0.91-1.11) | 0.90 |
| **Black female vs Black male (reference)** |  |  |
| Overall | 1.13 (1.03-1.24) | 0.009 |
| ATTRwt-CM | 1.13 (1.03-1.24) | 0.009 |
| ATTRv-CM | 1.13 (1.03-1.24) | 0.009 |
| SDI ≤80 | 1.18 (1.06-1.31) | 0.002 |
| SDI >80 | 0.98 (0.87-1.10) | 0.73 |

ATTRv-CM = variant transthyretin amyloid cardiomyopathy; ATTRwt-CM = wild-type transthyretin amyloid cardiomyopathy; HR = hazard ratio; SDI = Social Deprivation Index.

**Supplemental Figure 1.**
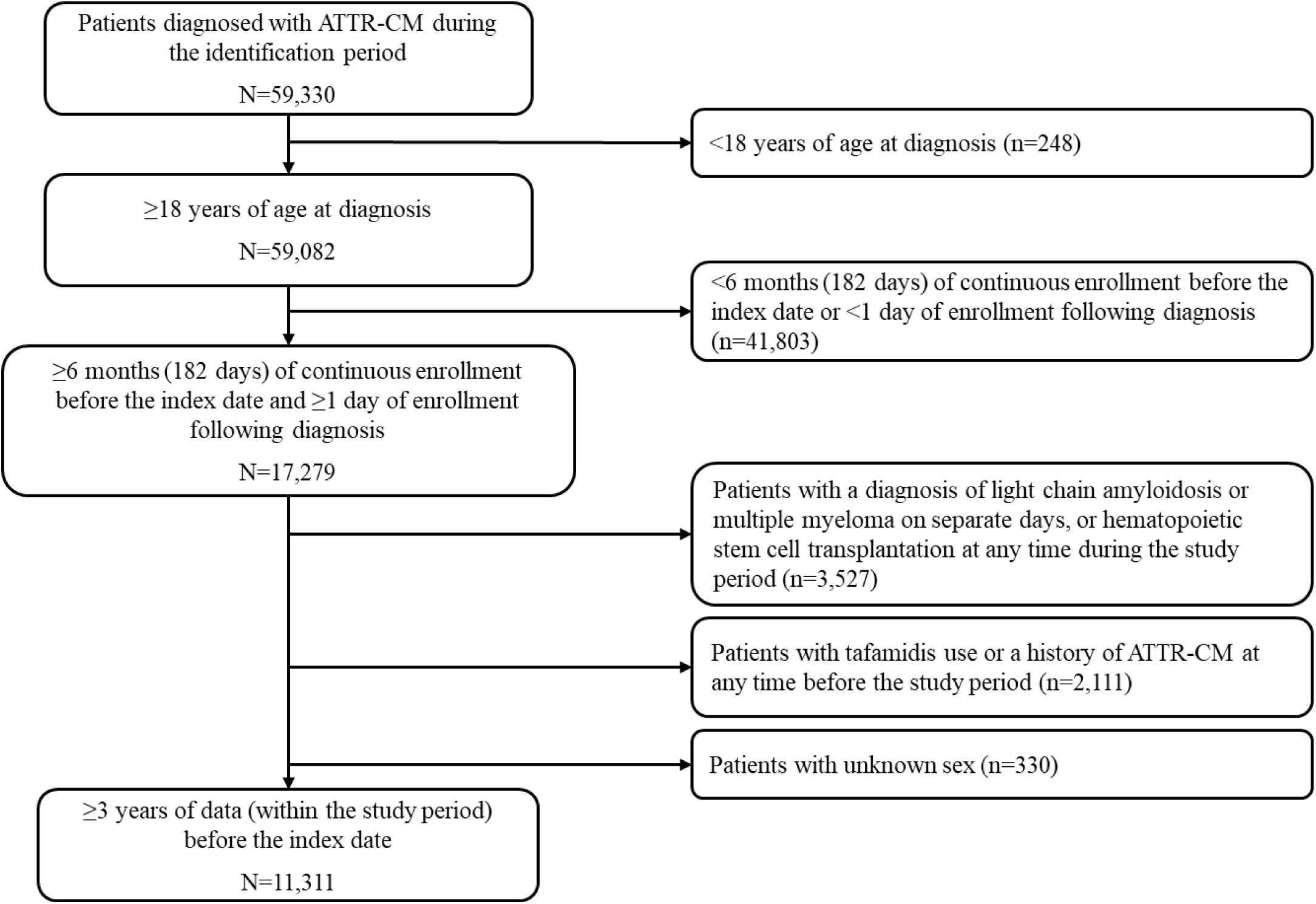
CONSORT Flow Diagram. ATTR-CM = transthyretin amyloid cardiomyopathy.

## Notes

### Author Declarations

The Komodo database complies with the Health Insurance Portability and Accountability Act. Given the deidentified nature of the data, patient consent and institutional review board approval were not required.

